# Reactivating Residual Motor Circuits in Complete Spinal Cord Injury Through Mode- and Site-Specific Transcranial Magnetic Stimulation: A Pilot Randomized Controlled Trial

**DOI:** 10.64898/2026.10.31.26366692

**Authors:** Deeksha Patel, Rohit Banerjee, Suman Jain, Kamran Farooque, Asjad Mahmood, Hanuman P Sharma, Deepak Gupta, Nand Kumar

## Abstract

**Background:** Recovery after motor complete spinal cord injury (cSCI) remains limited, though spared fibers and latent neural circuits may support neuroplasticity. Transcranial magnetic stimulation (rTMS) and intermittent theta burst stimulation (iTBS) induces LTP-like plasticity, however their comparative efficacy in cSCI remains unclear. We compared rTMS/ iTBS; iTBS at various sites combined with rehabilitation on motor outcomes and their underlying mechanism following cSCI.

**Methods:** In this double-blind randomized controlled trial, 37 individuals with thoracolumbar AIS-A (<3 months post-injury) were randomized to five groups: sham, motor-cortex rTMS, motor-cortex iTBS, spinal-cord iTBS, and combined motor-cortex–spinal-cord iTBS. 30 participants who completed follow-up (n=6/group), received 2 TMS sessions/day X 5 days alongside standardized rehabilitation. Lower Extremity Motor Score (LEMS) was the primary outcome; secondary outcomes included functional, electrophysiological, biochemical, neurotransmitter, pain and quality-of-life measures, assessed at baseline, post-intervention, and two follow-ups.

**Results and Conclusion:** Both iTBS-MC and iTBS-MC+SC groups showed an increase in LEMS and improved WISCI-II scores (p < 0.05). LEMS strongly correlated with WISCI (R = 0.60, p = 0.0019), suggesting the regain of assisted ambulation by FU2. ΔLEMS showed positive correlation with MEP_amplitude_ (R = 0.49, p = 0.014), area under the curve (AUC; R = 0.43, p = 0.034) and negative correlation with pNF-H (R = −0.54, p = 0.0006), indicating enhanced recruitment of dormant pathways and reduced neuronal injury. Glutamate levels also negatively correlated with ΔLEMS (R = -0.4, p = 0.05). Quality of life improved significantly (p< 0.05) in both rTMS and iTBS groups.

**Clinical Trial Registry India:** CTRI /2022/11/047038

## 1. Introduction

Complete spinal cord injury (cSCI) causes profound disruption of descending motor and ascending sensory pathways, leading to long term functional deficits. Despite advances in acute care and rehabilitation, recovery has remained limited in these individuals clinically classified as complete injury [1]. For decades, the absence of voluntary motor and sensory function below the level of injury was interpreted as evidence of complete anatomical disruption of spinal pathways, resulting in the belief that functional restoration beyond a brief period of spontaneous recovery was unlikely [2]. However, clinical observations and neurophysiological studies involving recordings of motor (MEPs) and somatosensory evoked potentials (SSEPs) in SCI provided evidence for preservation of feeble signal transmission across the lesion site, suggesting presence of anatomically intact but functionally inactive pathways [3,4]. This evolving perspective shifted the SCI research beyond structural preservation to restoration of spared and latent neural connections by neuromodulation strategies. The present mechanistic study addressed this initiative by exploring the therapeutic potential of Transcranial magnetic stimulation (TMS) along with standardized physical rehabilitation on inter and intra-cortical plasticity and excitability

Conventional high-frequency repetitive TMS (rTMS) has been shown to enhance cortical excitability and improve sensorimotor outcomes in incomplete SCI; its effect on motor recovery remain inconsistent in cSCI [5,6]. Intermittent theta burst stimulation (iTBS), a patterned form of TMS, is a more efficient alternative; engaging mechanisms similar to long-term potentiation (LTP) and inducing robust and sustained plastic changes within cortical networks [7,8]. iTBS is shown to modulate corticospinal excitability and facilitates neuroplasticity-related signaling pathways by BDNF-mediated mechanisms [9,10]. However, its clinical efficacy and comparative effectiveness with rTMS remains to be studied in cSCI.

The anatomical stimulation site can critically influence therapeutic outcomes. Most studies in SCI have targeted primary motor cortex (M1), given its central role in voluntary motor control. However, motor function depends on coordinated interactions between supraspinal and spinal networks, and modulation at the level of the spinal cord can engage segmental circuits and afferent pathways not directly accessible through cortical stimulation alone [11,12]. The brain–spine interface studies suggests that combined cortical and spinal stimulation can enhance recovery through synergistic mechanisms [13,14]. So, the contributions of stimulation site—cortical or spinal or combined—needs to be systematically evaluated.

Interpretation of neuromodulation data is further limited by the lack of integrated mechanistic assessment. Clinical outcomes provide evidence of efficacy but do not fully capture the underlying biological processes. Neurophysiological measures: MEPs and recruitment curve parameters, paired pulse TMS paradigms offer insights into corticospinal excitability and intracortical excitatory-inhibitory networks. Additionally, circulating biomarkers related to neuroplasticity, inflammation, and axonal injury provides complementary information on molecular processes underlying recovery [15]. These domains are often examined in isolation, limiting the ability to link physiological modulation with functional improvement.

In the present study, a unified comparative, site-specific design was implemented with longitudinal assessment across clinical, electrophysiological, biochemical, and psychosocial domains, incorporating correlation-based analyses to link functional outcomes with biological changes and assess re-engagement of dormant pathways. The objectives were to compare the efficacy of different (1) stimulation modes and (2) anatomical target sites, as well as to establish their integrated mechanistic correlates in complete SCI.

## 2. Materials and Methods

### 2.1 Study Design and Participants

This randomised, double-blind, placebo-controlled trial compared the efficacy of different (1) stimulation modes (Placebo vs rTMS vs iTBS) over the M1 (MC); and (2) anatomical sites using iTBS (MC or spinal cord (SC) or combined (MC+SC). The details of the study design and protocol as elaborated in Patel et al., (2025) was followed [16].

30 participants with acute traumatic thoracolumbar SCI (T1–L2), AIS grade A, aged 18–60 years, enrolled within three months of injury, were randomised into five groups (n = 6 per group): Placebo, rTMS-MC, iTBS-MC, iTBS-SC, and iTBS-MC+SC. Exclusion criteria included patients with AIS grade B–D, tetraplegia, epilepsy, metallic implants, pressure ulcers, fractures, and inability to participate in rehabilitation program.

Ethics approval was obtained from Institutional Ethics Committee (IEC no. IECPG-551/28.07.2022). The study was conducted in accordance with the Declaration of Helsinki and written informed consent was obtained from all participants. The trial was registered at Clinical Trials Registry – India with registration no. CTRI/2022/11/047038. The trial was conducted between November 2022 to February 2026. The patients were recruited from department of Orthopaedics, Trauma Centre, All India Institute of Medical Sciences (AIIMS), Delhi. The baseline assessments and interventions were given at Brain Stimulation laboratory, department of Physiology, AIIMS, Delhi.

#### Patient and public involvement

Patients and members of the public were not involved in the design, conduct, reporting, or dissemination plans of this research. Participants were involved as study participants and provided written informed consent before enrolment.

### 2.2 Patient Recruitment, Randomisation, and Replacement Protocol

83 SCI patients were screened, 46 were excluded (not meeting inclusion criteria) and 37 were randomised (Figure_1). Randomisation used a computer-generated concealed allocation sequence. Participants and outcome assessors were blinded; the TMS operator was unblinded to protocol and thus excluded from assessment.

**Figure_1.**
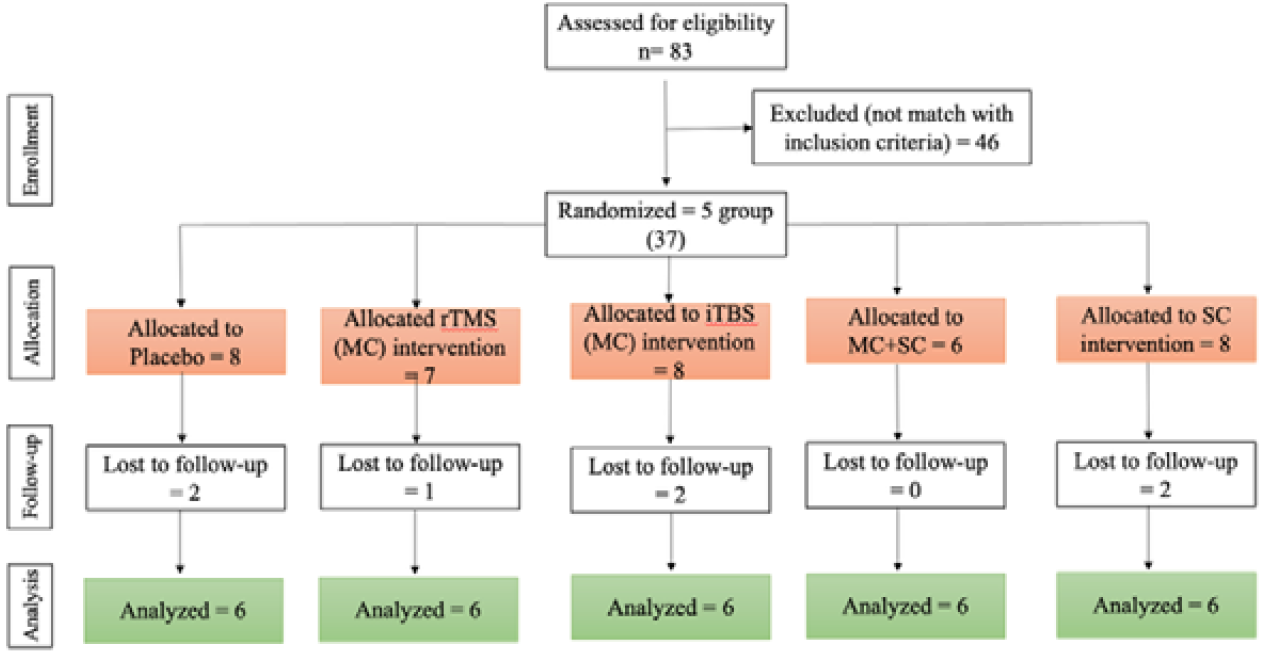
CONSORT diagram.

Given the small target sample (n = 6 per group), a pre-specified replacement recruitment protocol was implemented: any participant unable to complete all sessions and assessments, was immediately withdrawn and replaced by the next eligible recruited patient within the same group, prior to analysis. Seven participants were withdrawn during the study (Placebo: n = 2; rTMS-MC: n = 1; iTBS-MC: n = 2; iTBS-SC: n = 2) due to loss of contact. No adverse events were recorded during or after intervention.

### 2.3 Placebo and Active Intervention

rTMS or iTBS was delivered at 90% RMT (Neuro-MS 5 system, Neurosoft Ltd®, Ivanovo, Russia) over the leg area of the M1, 2 sessions/ day (at minimum 2-hour interval) over 5 consecutive days, for a total of 10 sessions. Each session was followed by standardised rehabilitation. Placebo stimulation protocol was similar to active but without any magnetic pulses (Figure_2).(details in *Supplementary Methods S1*).

**Figure_2.**
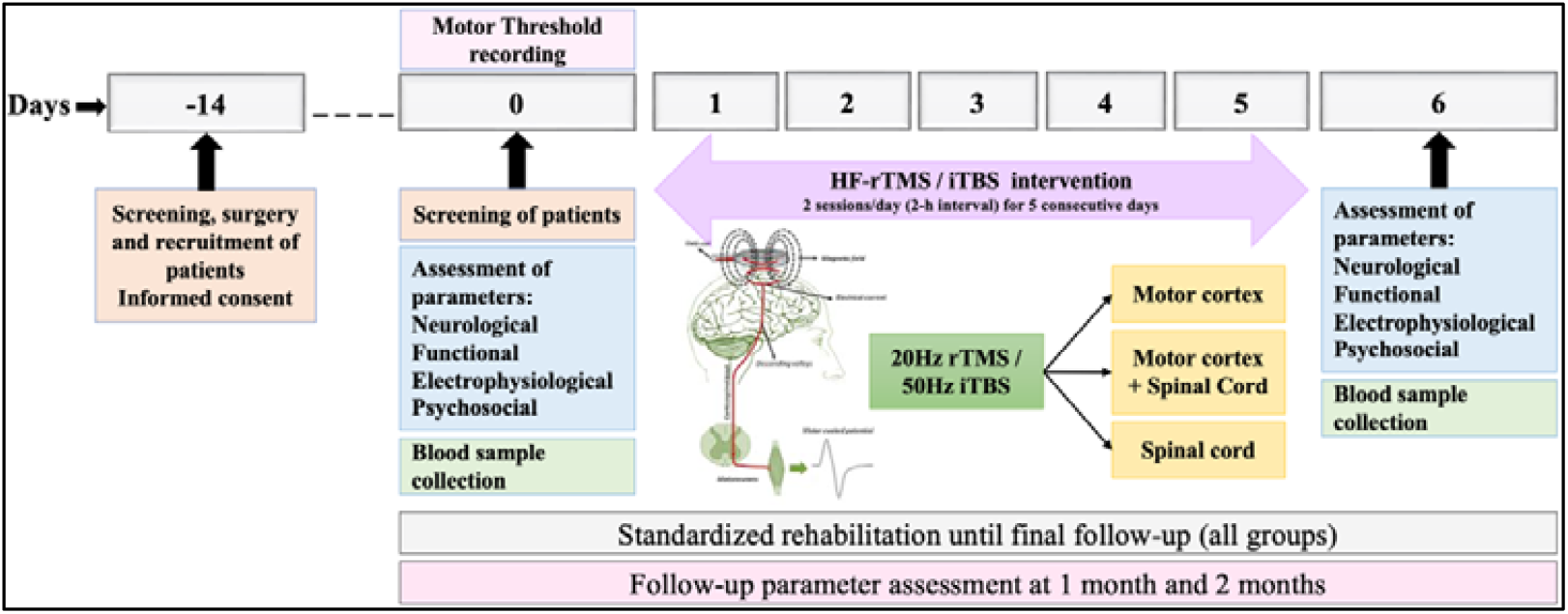
Schematic representation of Experimental design

### 2.4 Clinical, Electrophysiological and Biochemical Outcomes

All clinical and electrophysiological assessments were conducted at baseline (Pre), post-intervention (Post; day 6), and at one (FU1) and two-month (FU2) follow-ups. The AIS Score included lower extremity motor score (LEMS; 0–50) and sensory function scoring, with LEMS as the primary outcome. Secondary outcomes included WISCI-II, SCIM-III (self-care, respiration/sphincter management, mobility), TMS single-pulse parameters (RMT, MEP recruitment curve, cSP), TMS paired-pulse parameters (SICI, ICF, LICI), VAS (pain), BDI-II, STAI (Y1/Y2), and WHOQOL-BREF (*Supplementary Methods S2, S3, S5)*.

Blood samples were collected for biochemical analysis at Pre, Post, and at FU2, centrifuged and stored at −80°C. Serum BDNF, NT-3, pNF-H, MBP, IL-6, and CD95L were quantified by sandwich ELISA in duplicate. Plasma GABA and glutamate levels were measured only in Objective 2 by LC-MS/MS with isotope-labelled internal standards (*Supplementary Methods S4)*

#### Statistical Analysis

Analysis were performed using R (v4.4.0; lme4, lmerTest, emmeans) and SPSS (v25.0) software. Total 30 patients (n = 6 per group) were analysed at all time points. Primary longitudinal analysis used linear mixed-effects models (LMM) with fixed effects for calculating group, time, group × time interaction, random participant intercept and degrees of freedom (Satterthwaite’s method). Significant interactions were followed by pairwise post-hoc evaluation using estimated marginal means with Tukey correction (within-group temporal analysis; between-group: comparison of groups at each time point). Non-parametric sensitivity analysis used the Friedman test (within-group) and Kruskal–Wallis with Dunn’s test (between-group) for post-hoc comparisons. Biomarker data were log-transformed for analysis and FDR correction (Benjamini–Hochberg) applied across biomarker outcomes. Effect sizes were quantified using Kendall’s W (Friedman) and epsilon squared (Kruskal– Wallis) tests. All correlations were performed using Spearman’s rank correlation coefficient (two-tailed) on change scores (Δ = Follow-up minus Baseline). All tests were two-tailed (α = 0.05); exact p-values are reported with adjustment method indicated.

## 3. Results

### Participant characteristics and baseline comparison

Baseline data for demographic and injury characteristics; clinical outcomes; level of serum biomarkers and psychosocial outcomes were comparable across groups. No significant differences were observed among the groups for age, gender, time since injury or AIS score (p ≥ 0.05) *(Supplementary Table S1)*.

#### Objective 1: Effect of Stimulation Mode (rTMS vs iTBS)

##### Primary outcome: Lower extremity motor score

LEMS demonstrated a significant time X group interaction in the iTBS group at FU2 (p = 0.018). It increased significantly following iTBS intervention, from baseline to the 2^nd^ FU (p = 0.0013) (Figure 3a, 3b). No significant between-group difference was evident.

**Figure_3.**
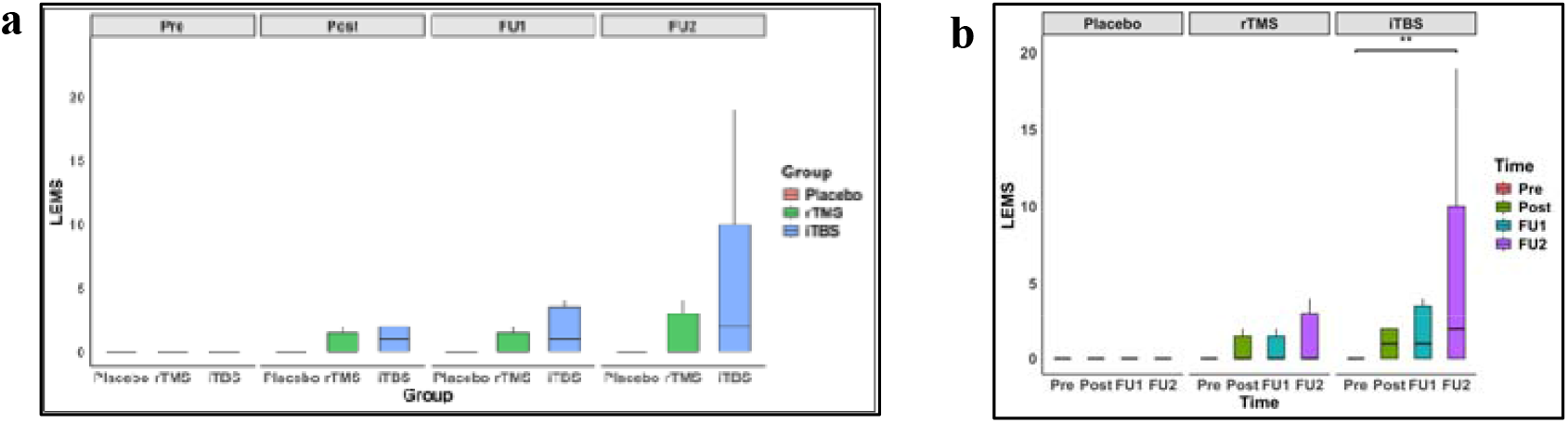
Lower extremity motor score (LEMS) following rTMS or iTBS intervention. LEMS score (median with interquartile range) was evaluated across four time points: pre-intervention (Pre), post-intervention (Post), first follow-up (FU1), and second follow-up (FU2). (a) between-group comparisons. (b) within-group comparisons. **p < 0.01

AIS sensory score analysis is given in Supplementary material *(Figure S1a, S1b* and *Table S2)*.

##### Secondary outcomes: a) Functional outcome

*Walking Index for Spinal Cord Injury (WISCI)* score was significantly higher following iTBS at FU1 and FU2, in comparison to both placebo (p = 0.001; p = 0.020) and rTMS (p = 0.033; p = 0.012) groups. At FU2, rTMS intervention also showed higher scores than placebo (p = 0.011). Further, as compared to baseline, iTBS intervention significantly increased in WISCI (p < 0.001) at FU1 and FU2, while rTMS showed significant increase only at FU2 (p < 0.001) (Figure 4a, 4b).

**Figure_4.**
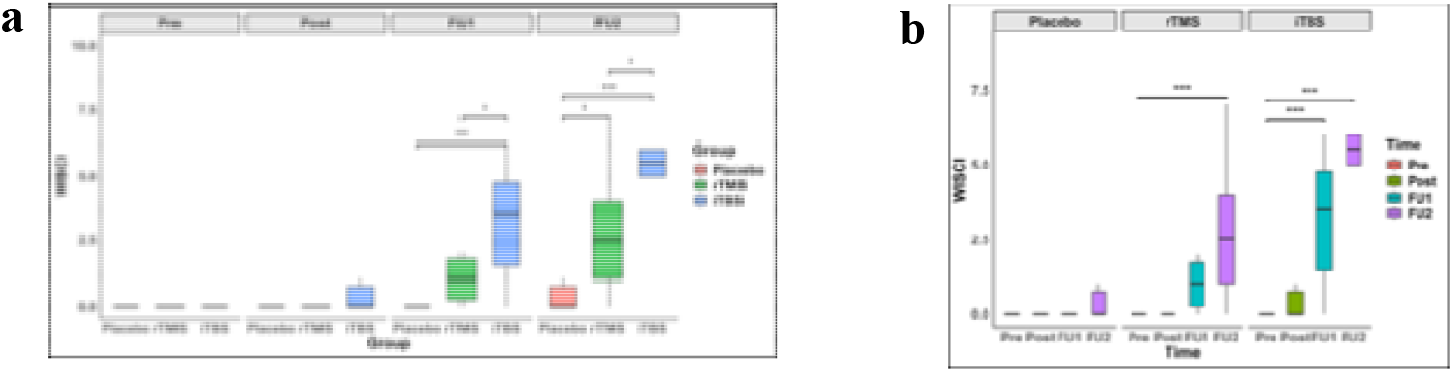
Walking index for SCI (WISCI) (score; Median with interquartile range) across stimulation modes (a) intergroup comparisons at each time point and (b) within-group comparison over the time points. *p < 0.05, **p < 0.01, ***p < 0.001.

###### Functional independence

SCIM domain scores (self-care, respiration and sphincter management, and mobility) complemented the WISCI findings (*Supplementary material and result, Figure S2*; *Table S2*).

##### b) Electrophysiological outcomes

At baseline, all three groups showed overlapping, sigmoidal *MEP recruitment curves* with comparable MEP_max_ (∼1–2 mV) and I_50_ (∼75–80% MSO). A significant effect of both the interventions was evident by a steep increase in the amplitude with progressive increase in threshold intensity. However, iTBS intervention led to an increased amplitude at an earlier time point (Post-intervention) as well as at lower intensities as evident by a leftward shift of the curve (Figure 5a). AUC, MEP_max_ and I_50_, all demonstrated a significant group, time, and time x group interaction effects (p < 0.01) (details in *Supplementary results, Figure S3*; *Table S2*).

**Figure_5.**
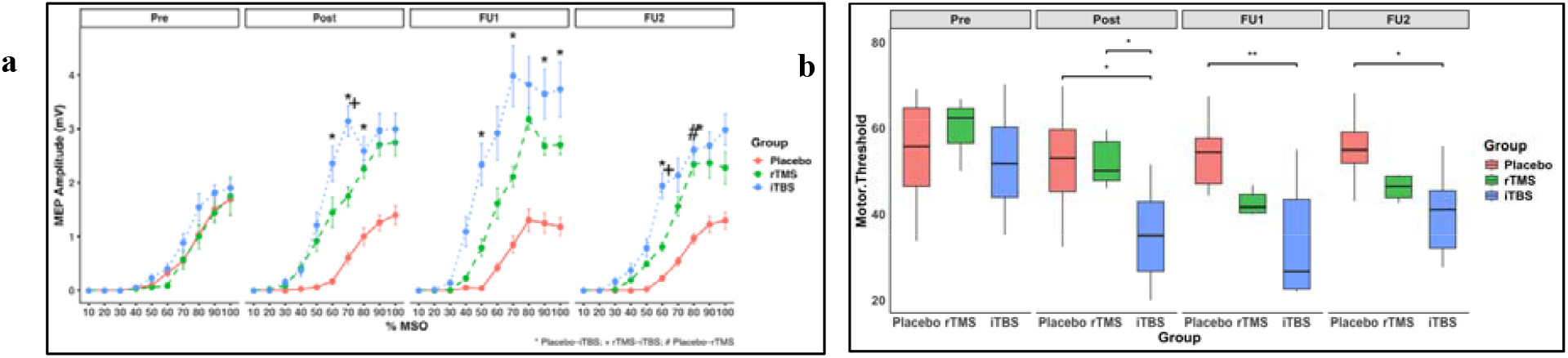
MEP recruitment curves and motor threshold in various groups. (a) MEP recruitment curves (amplitude (mV); Mean ± SEM)) at four time points: pre-intervention (Pre), post-intervention (Post), first follow-up (FU1), and second follow-up (FU2). *p < 0.05 (Placebo vs iTBS), +p < 0.05 (rTMS vs iTBS), #p < 0.05 (Placebo vs rTMS). **(b)** Motor threshold (% MSO, Median with interquartile range) at four time points *^/+/#^p < 0.05, **p < 0.01.

*Motor threshold* was significantly reduced in iTBS group as compared to placebo across all time points (Post: p = 0.019; FU1: p = 0.004; FU2: p = 0.039) and compared to rTMS at post-intervention (p = 0.019) (Figure 5b).

*Cortical silent period (CSP)*, did not show statistically significant difference among the groups. (*Supplementary Figure S4; Table S1; Table S2)*.

##### c) Biochemical outcomes

A significant time X group effect was observed, with iTBS showing significant increase in serum BDNF, NT-3 and MBP as compared to placebo (p ≤ 0.05) (*Supplementary results, Supplementary Figure S5; Table S1; Table S2*).

##### d) Psychosocial outcomes

A significant effect of iTBS intervention was observed on psychosocial outcomes as evident by a significant increase in WHO-QOL Physical and Psychological domain scores at FU2 as compared to placebo (p ≤ 0.05). BDI and VAS scores decreased significantly (p ≤ 0.05) as compared to baseline in both rTMS and iTBS (*Supplementary results, Figure S6; Table S2*).

### Correlation analysis

ΔLEMS showed a significant positive correlation with ΔWISCI at FU2 (R = 0.68, p = 0.002). ΔWISCI correlated positively with the physical subdomain of WHOQOL, ΔMEP, ΔAUC, ΔNT3 and mobility subdomain of SCIM (*Supplementary results, Figure S7)*.

#### Objective 2: Effect of iTBS Stimulation Site on primary and secondary outcomes

##### Primary outcome: Lower extremity motor score and sensory score

A significant time X group interaction was observed in LEMS following iTBS intervention on M1 (MC) at FU2 (p = 0.008). Post hoc analysis showed a significant increase of LEMS in both iTBS (MC) and iTBS (MC+SC) groups at FU2 (p = 0.0002; p = 0.009 respectively) as compared to baseline. Further, LEMS showed facilitation in iTBS(MC+SC) group as compared to placebo at FU2 (p = 0.040) (Figure 6a, b).

**Figure_6.**
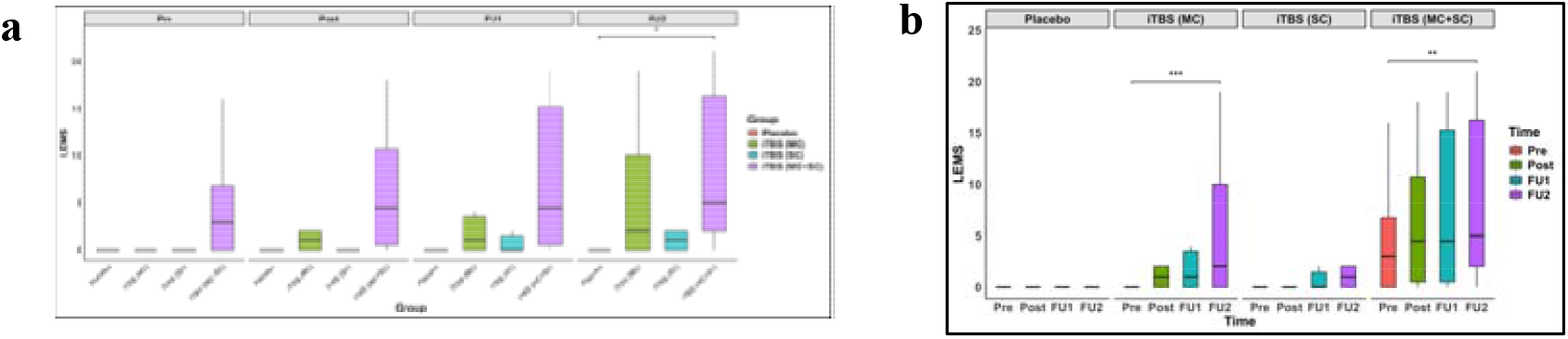
Lower extremity motor scores. (Median with interquartile range) (a) comparisons of LEMS across Placebo, iTBS(MC), iTBS(MC+SC) and iTBS(SC) groups at each time point. (b) depict within-group temporal comparisons. *p < 0.05, **p < 0.01, ***p < 0.001.

Sensory function also showed significant effect of iTBS intervention on MC in comparison to placebo at FU2 (p<0.05) (*Supplementary material, Figure S8, Table S3*).

##### Secondary outcomes: a) Functional outcome

*Walking Index for Spinal Cord Injury (WISCI)* increased significantly at FU1 and FU2 as compared to placebo, in both iTBS(MC) (p = 0.023; p = 0.0002) and iTBS(MC+SC) (p = 0.0002; p < 0.0001) groups. Additionally, WISCI was also significantly more in iTBS (MC+SC) as compared to iTBS (SC) at both FU1 and FU2 (p = 0.001; p < 0.0001) (Figure 7a). Within-group analysis found significant increase in both iTBS(MC) and iTBS(MC+SC) groups at FU1 and FU2 as compared to baseline (p < 0.01) (Figure 7b).

**Figure_7.**
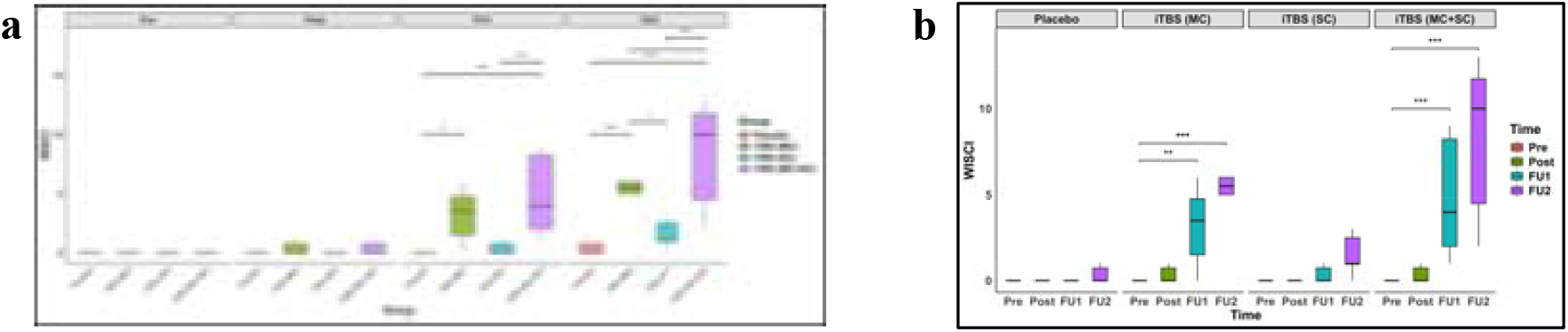
Walking index for SCI. (score; median with interquartile range) (a) between-group comparisons across Placebo, iTBS(MC), iTBS(MC+SC) and iTBS(SC) groups at each time point and (b) within-group temporal comparisons. *p < 0.05, **p < 0.01, ***p < 0.001.

###### Functional independence

A significant improvement in SCIM domain scores (self-care, respiration and sphincter management, and mobility) was evident in iTBS (MC), and iTBS (MC+SC) groups at follow-ups as compared to Placebo as well as from baseline (*Supplementary Figure S9, Table S1, S3*).

##### b) Electrophysiological outcomes

###### MEP Recruitment Curve

At baseline, all four groups showed overlapping recruitment curves (MEP_max_ ∼1 mV and I_50_ ∼75-80% MSO). A significant effect of iTBS stimulation at MC as well as at combined MC and SC was observed. Both these {iTBS(MC) and iTBS(MC+SC)} groups showed pronounced leftward shifts with substantially elevated plateaus at Post, FU1 and FU2 (MEP_max_ ∼3–4.5 mV and I_50_ ∼50–55% MSO) (Figure 8a). AUC in these groups was also significantly (p ≤ 0.001) higher at all time points as compared to iTBS (SC) and baseline. MEP_max_ increased significantly at FU1 (p = 0.035) and I_50_ showed a decrease at Post (p = 0.0009) in iTBS(MC) group as compared to placebo (Figure 8b; *Supplementary Figure S10, Table S1, S3)*.

**Figure_8.**
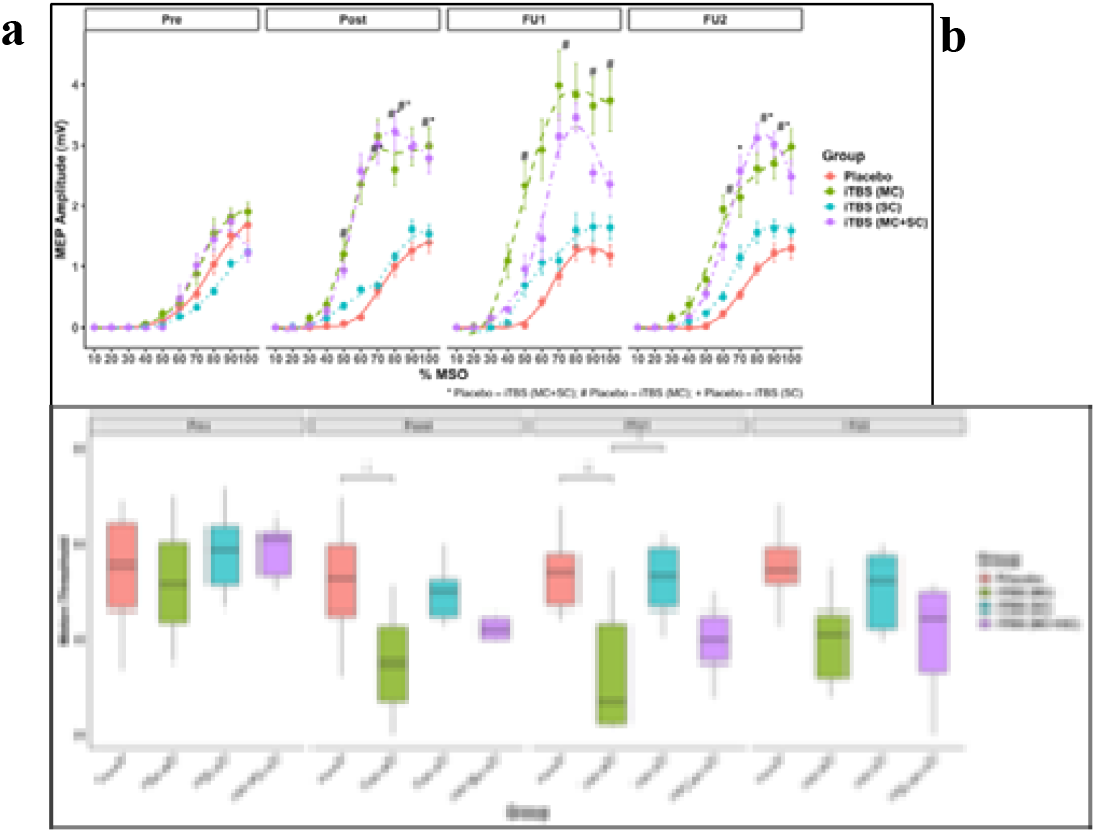
MEP recruitment curves and Resting Motor Threshold (RMT) **(a)** MEP amplitude (mV, Mean ± SEM) plotted against stimulation intensity (%MSO) across four time points for each group **(b)** RMT (%MSO, Median with interquartile range) of various groups. * (Placebo-iTBS (MC+SC), # (Placebo-iTBS (MC), + (Placebo-iTBS (SC). */#/+p < 0.05, **p < 0.01.

*Resting Motor Threshold* in iTBS(MC) was found to be significantly lower than placebo at Post (p = 0.040) and FU1 (p = 0.009) (Figure 6b). *Cortical silent period* did not show difference among the groups (*Supplementary material, Figure S11, Table S2)*.

###### SICI-ICF

At baseline, all four groups showed typical paired-pulse response, with overlapping input-output curves of SICI-ICF and % MEP below 100%, suggesting overall inhibition in all the groups. However, following intervention at follow up periods, the iTBS-MC and iTBS-MC+SC groups showed a clear leftward and upward shift in the IOC curve during 10–30 ms, suggesting progressively higher conditioned MEP amplitudes at the longer ISIs (Figure 9a). At FU2, in all the iTBS-MC, iTBS-SC and iTBS-MC+SC groups significantly higher SICI values were observed than the placebo group (*p* ≤ 0.001). ICF (7-30msec) was significantly (p < 0.001) increased in iTBS(MC) and iTBS(MC+SC) groups at all time points post intervention as compared to placebo. Additionally, iTBS(MC+SC) also showed significant increase in ICF as compared to iTBS (SC) group (*p* ≤ 0.01) (*Supplementary results, Figure S12*).

**Figure_9.**
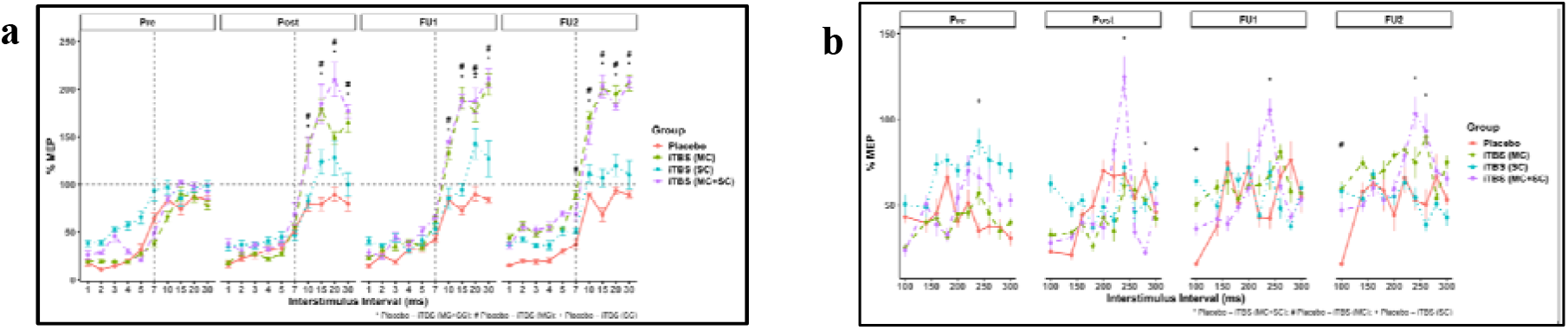
Modulation of intracortical inhibition and facilitation across stimulation sites. (a) short-interval intracortical inhibition (SICI; 1–5 ms) and intracortical facilitation (ICF; 7–30 ms), expressed as %MEP (Mean ±SEM) across interstimulus intervals at four time points; (b) long-interval intracortical inhibition (LICI; 100–300 ms) across the same time points. Data are presented for each group (Placebo, iTBS (MC), iTBS (SC), iTBS (MC+SC)). Dashed horizontal line represents baseline (100% MEP). (*Placebo-iTBS (MC+SC), #Placebo-iTBS (MC), + Placebo-iTBS (SC); p < 0.05.

###### LICI

IOC curves of all four groups largely overlapped at baseline and Post-intervention. However, in the iTBS-MC and iTBS-MC+SC groups higher conditioned MEP responses were observed at the longer ISIs during the follow-ups (Figure 9b). At FU2, both the iTBS-MC and iTBS-MC+SC groups showed significant increase in % MEP than the placebo group (*p* = 0.005 and *p* = 0.046 respectively). Within group analysis also revealed significant increase in both these groups (*Supplementary results, Figure S13*).

##### c) Biochemical outcomes

BDNF was significantly higher at follow-up in both iTBS(MC) and iTBS(MC+SC) as compared to iTBS (SC) group (p < 0.001). Glutamate levels also significantly increased at follow-up in iTBS(MC) in comparison to placebo p < 0.001 and iTBS(SC) p < 0.001. However, pNF levels decreased significantly in iTBS (MC) as compared to iTBS(SC) (*Figure S14, Table S1, S3*).

##### d) Psychosocial outcomes

WHO-QOL Physical and Psychological, domain scores increased significantly (p < 0.001) in iTBS(MC+SC) as compared to placebo at follow-ups. Within group comparisons over the time points also showed a significant increase in all domains. BDI and VAS showed significant within-group reductions across all active groups (*Supplementary Figure S15 , Table S1, S3*).

### Correlation analysis

ΔLEMS was positively correlated with ΔWISCI (R = 0.60, p = 0.002), ΔMEP amplitude,

ΔAUC and negatively correlated with Δ pNF at FU2 (*Supplementary results, Figure S16)*.

## 4. Discussion

To the best of our knowledge, this is a novel randomised controlled, double-blind trial sequentially comparing the therapeutic potential of two different TMS protocols (rTMS vs iTBS) on motor recovery and evaluating optimum site for iTBS in individuals with complete SCI. The results provided evidence of iTBS producing meaningful improvement in lower limb muscle strength and initiation of stepping. Functional recovery was specifically augmented by combined stimulation of the motor cortex and spinal cord. Further, the motor recovery correlated with changes in both electrophysiological parameters and circulating biomarkers. Administration of rTMS, though showed functional recovery with time, it was significantly less than iTBS. The more pronounced effect of iTBS is likely due to its theta-burst delivery pattern, which is thought to effectively engage LTP-like synaptic mechanisms that strengthen residual corticospinal pathways and promote targeted cortical reorganization compared with conventional rTMS [17].

Motor score, as indexed by LEMS showed a delayed but progressive significant increase at 2^nd^ follow-up as compared to placebo, in combined iTBS group. This improvement in muscle strength over time suggests that recruitment of surviving neurons is not an immediate result of stimulation, but gradual activation of spared neurons. Spinal stimulation increases the excitability of lumbosacral neurons, which progressively activates the residual functionally preserved neural circuits below the level of lesion. Cortical stimulation during this time, in turn activates the descending corticospinal pathways. Thus combined stimulation steadily strengthens the communication within the sensorimotor network, resulting in greater neurological recovery than that achieved with cortical or spinal stimulation alone [18-19]. Walking ability (WISCI), RSM and mobility subdomain of SCIM was increased in the iTBS (MC) and iTBS (MC+SC) groups even at the first follow-up as compared with placebo. Early gains in functional outcomes may be due to the enhancement in recruitment of spinal locomotor circuits, including central pattern generators (CPGs), enabling better stepping and mobility even before substantial improvements in muscle strength are manifested [20-21]. Within group improvement in self-care subdomain of SCIM was evident in all groups, including placebo, suggesting substantial role of extensive rehabilitation provided to individuals twice daily; training induced neuroplasticity [22-24]. Thus, iTBS administration seems to strengthen the communication between the brain and spinal motor circuits by amplifying the response of rehabilitation. iTBS-SC group showed an indistinguishable recovery as compared to placebo, indicating that spinal cord stimulation alone may be inadequate to promote motor recovery after acute SCI. The increase in walking capacity also led to improved quality of life as evident by enhanced scores of physical and psychological WHO-QOL subdomains [25]. Unlike motor recovery, sensory function; light touch and pin prick scores, improved in both rTMS and iTBS groups, which can be partly explained by the relatively large size of the figure-of-eight coil. Owing to the close anatomical proximity of the primary motor and somatosensory cortices, the magnetic field generated by the coil is likely to stimulate both regions, thereby promoting sensory recovery [26].

Recruitment curves (RC) or TMS input–output (IO) curves are extensively used as a measure of cortical plasticity and corticospinal excitability [27]. It assesses the relationship between the amplitude of MEP evoked in the target muscle and single pulse TMS stimulation intensity. The steepness of the curve (slope), midpoint value I_50_ and MEP_max_ are considered to be an independent and reliable metrics of neuroplasticity [28]. The steeper IO slopes and lower I_50_ values correlate positively with MEP facilitation [29-31], thereby the presence of a highly responsive corticospinal circuit. In our study, RC analysis demonstrated an increase in MEP_max_ and a reduction in I_50_ following iTBS-MC, indicating increased excitability and responsiveness of corticospinal neurons. In iTBS, repeated bursts delivered at a theta frequency reduce feed-forward inhibition and promote greater postsynaptic depolarization, thereby facilitating NMDA receptor activation and enhancing induction of LTP. This increases the efficiency of synaptic transmission and responsiveness of corticospinal neurons to produce more sustained changes in cortical excitability as evident by a significant reduction in RMT [32, 33]. Among all recruitment curve parameters, AUC demonstrated the most consistent changes, increasing significantly after post-intervention in both iTBS-MC and iTBS-MC+SC and remained elevated throughout the follow-up periods. As AUC reflects the cumulative motor response across the entire range of stimulation intensities, its sustained increase suggests a persistent enhancement in the pattern of motor unit recruitment. Generally, at low TMS intensity, electrophysiological recordings show I1-waves, whereas, at higher intensities, it becomes complex and there is recruitment of later indirect waves (I2–I4 waves), which are generated via trans-synaptic activation of pyramidal neurons in the motor cortex by excitatory neurons. At higher intensities, there is cumulative recruitment and activation of inactive and silent neurons in both local and distant cortical circuits [34], contributing to increased AUC.

Paired-pulse TMS measures provided additional insight into the underlying neurophysiological mechanisms. As cortical excitability is governed by a complex interplay of excitatory and inhibitory neuronal circuitry impinging upon pyramidal neurons, ppTMS provides a comprehensive reliable tool to assess the relative physiological contribution of each of them [35]. While SICI is due to activation of GABA_A_ receptors inhibitory interneuronal cells, LICI activates GABA_B_ receptors and ICF increases the excitability in a NMDA receptor dependent manner [36]. SICI was significantly reduced at the second follow-up, whereas ICF increased immediately after the intervention and remained elevated throughout the follow-up periods in both the iTBS-MC and iTBS-MC+SC groups. In addition, LICI was significantly reduced at the final follow-up, indicating that iTBS modulated both GABA A and B type of receptors. iTBS has shown to alter the parvalbumin and calbindin expression, influencing intracellular calcium buffering and calcium-dependent neurotransmitter release in distinct GABAergic interneuron populations. Reduced parvalbumin-mediated perisomatic inhibition and altered calbindin-dependent dendritic regulation may shift the excitation–inhibition balance toward enhanced glutamatergic transmission and reduced GABAergic inhibitory control, facilitating synaptic plasticity and corticospinal pathway activation [37].

Importantly, WISCI showed a significant positive correlation with ICF and a negative correlation with SICI, indicating that greater intracortical facilitation and reduced intracortical inhibition is associated with better walking ability. These correlations indicate that ability to walk is associated with an optimum balance between intracortical facilitation and inhibition circuitry, that was restored by iTBS administration.

A reduction in circulating glutamate during post-intervention in iTBS-MC+SC group and at FU in iTBS-MC was evident. An injury to the blood–spinal cord barrier (BSCB) permits greater exchange of glutamate between the systemic circulation and the CNS, forming blood-to-CNS concentration gradient, which promotes movement of glutamate out of the CNS, thereby reducing CSF glutamate and excitotoxic stress [38]. Serum GABA also decreased following interventions that reflect alterations in systemic GABA metabolism after injury and neuromodulation [39]. Interestingly serum GABA showed a significant correlation with WISCI. The mechanism of systemic GABA in promoting functional recovery is still elusive. However, these novel observations lay a foundation for future studies measuring GABA simultaneously in blood and CNS to determine how circulating GABA changes relate to functional recovery after SCI.

A significant increase in the levels of neuroplasticity markers was evident; BDNF, and NT-3 was increased in both iTBS-MC and iTBS-MC+SC groups at FU, while NT-3 also improved in later group at post intervention, suggesting its positive beneficial neurotrophic effect especially in the combined iTBS group. These findings align with the concept that activity-dependent plasticity is mediated by neurotrophins signaling pathways, particularly BDNF, which plays a central role in synaptic strengthening and motor learning [40]. The observed reduction in serum MBP hints towards attenuated demyelination along with beneficial effect of stimulation on myelin remodeling or stabilization, consistent with evidence that neural activity regulates adaptive myelination and oligodendrocyte function [41]. The reduction in IL-6 across all groups, including placebo, suggest that inflammatory signaling is likely affected by rehabilitation or natural recovery rather than neuromodulation [42].

A key strength of this study lies in its integrated and novel design, being among the first to systematically combine both stimulation mode and site within a single randomized framework, while simultaneously evaluating their effects across neurological, functional, electrophysiological and biochemical domains to provide a mechanistic understanding of recovery. However, these strengths should be interpreted in light of certain limitations. The sample size was relatively small, potentially limiting detection of subtle between-group differences. Paired-pulse measures were available only in a subset due to mid-study equipment acquisition. Stimulation intensity was based on upper limb thresholds and may not fully reflect lower limb circuitry. The follow-up duration was limited, precluding long-term assessment. Future studies should focus on larger, multicentric cohorts with extended follow-up and incorporate advanced neuroimaging or connectivity-based approaches to further delineate the network-level mechanisms and optimize individualized neuromodulation strategies.

## 5. Conclusion

These findings support a multidimensional treatment protocol in which recovery arises from the interaction of stimulation waveform and site along with extent of rehabilitation. iTBS enhances neuronal recruitment and rebalance intracortical excitatory-inhibitory networks, while biochemical changes create a permissive environment for neuronal plasticity in individuals with complete SCI. The combined stimulation seems to be more beneficial as stimulation of the motor cortex strengthens descending motor signals, while stimulation of the spinal cord makes spared local circuitry more responsive to those signals and rehabilitation helps these changes become stable, translating into functional improvement. The greater effects observed with combined stimulation also highlight the importance of engaging multiple neural networks. Neuromodulation, when combined with rehabilitation, can shift recovery from compensatory adaptation toward meaningful functional restoration. This integrated approach provides a foundation for developing more effective neurorehabilitation strategies after complete SCI.

## Supporting information

supplementary results

## Data Availability

All data produced in the present study are available upon reasonable request to the authors

## CRediT authorship contribution statement

**Deeksha Patel:** Writing – review & editing, Writing – original draft, Visualization, Validation, Methodology, Investigation, Formal analysis, Data curation, Conceptualization. **Rohit Banerjee:** Investigation, Data curation, Rehabilitation. **Suman Jain:** Writing – review & editing, Supervision, Software, Resources, Project administration, Funding acquisition, Software, Conceptualization. **Kamran Farooque:** Surgical intervention, Patient recruitment, Investigation, Visualization. **Asjad Mahmood:** Surgical intervention, Patient recruitment, Investigation. **Human P Sharma:** Validation, methodology, Biochemical analysis. **Deepak Gupta:**, Investigation, patient recruitment. **Nand Kumar:** Validation, Software, Data curation, Resources.

## Ethics declaration and consent to participate

Written informed consent to take part in the study and to publish the article has been obtained from all participants. The privacy rights of participants have been observed.

This study was performed in compliance with relevant laws, regulatory frameworks and guidelines where the research took place. This study was approved by the Institutional Ethics Committee (IEC no. IECPG-551/28.07.2022)

This clinical trial was registered with number CTRI /2022/11/047038 (ClinicalTrials.gov).

## Generative AI Declaration

No generative AI tools were used in the writing, editing, data analysis, or figure creation for this manuscript.

## Acknowledgements

We are grateful to all the patients who agreed to participate in the study, the clinicians who helped with patient recruitment, the Mr. Sanjeev Beniwal and Mr. Praveen Kumar delivering the intervention. We thank Mr. Ravi Verma for analysing the data.

---

**Supplementary Table S1:** Baseline analysis among the groups for Demographic and primary and secondary outcomes.

| Variable (Baseline) | Sham (n=6) | rTMS-MC (n=6) | iTBS-MC (n=6) | iTBS-SC (n=6) | iTBS-MC+SC (n=6) | p-value |
|---|---|---|---|---|---|---|
| Age (years) | 36.3 ± 7.87 | 31.7 ± 6.19 | 29.5 ± 8.14 | 32.3 ± 7.89 | 33.3 ± 7.71 | 0.80 |
| Sex (M/F) | 5/1 | 4/2 | 4/2 | 3/3 | 5/1 | 0.78 |
| Injury duration (months) | 1.83 ± 0.30 | 1.33 ± 0.21 | 1.50 ± 0.34 | 1.42 ± 0.34 | 1.58 ± 0.4 | 0.63 |
| AIS | A | A | A | A | A | - |
| LEMS | 0.0 ± 0.0 | 0.66 ± 1.63 | 1.33 ± 2.4 | 0.16 ± 0.4 | 2.6 ± 3.3 | 0.19 |
| WISCI | 0.0 ± 0.0 | 0.0 ± 0.0 | 0.0 ± 0.0 | 0.47 ± 1.12 | 0.0 ± 0.0 | 0.40 |
| SCIM | 19.1 ± 3.86 | 16.3 ± 2.87 | 18.3 ± 3.7 | 14.5 ± 4.18 | 17.5 ± 1.04 | 0.30 |
| BDI | 10.2 ± 1.8 | 11.1 ± 1.47 | 10 ± 2.16 | 11.2 ± 4.01 | 9.7 ± 3.50 | 0.69 |
| STAI | 96.8 ± 2.99 | 98.5.2 ± 4.5 | 100.8 ± 2.7 | 99 ± 3.22 | 98.3 ± 3.20 | 0.22 |
| VAS | 4.71 ± 1.23 | 4.52 ± 1.03 | 4.57 ± 1.20 | 4.52 ± 1.56 | 4.61 ± 1.24 | 0.93 |
| WHO-QOL | 70.1 ± 4.07 | 69.6 ± 2.58 | 69.1 ± 6.79 | 66.5 ± 2.25 | 65.1 ± 1.72 | 0.11 |
| MT (%) | 51.8 ± 9.61 | 60.1 ± 6.46 | 42.4 ±13.52 | 52.7 ± 9.0 | 46.1 ± 12.45 | 0.64 |
| cSP (ms) | 233 ± 64.6 | 233 ± 49.77 | 216 ± 53.4 | 198 ± 51.0 | 201 ± 51.8 | 0.59 |
| SICI | 19.7 ± 16.7 | - | 20.7 ± 9.8 | 27.3 ± 14.8 | 23.7 ± 16.08 | 0.06 |
| ICF | 66.4 ± 31.3 | - | 51.0 ± 33.6 | 54.9 ± 38.3 | 53.8 ± 35.3 | 0.41 |
| LICI | 40.9 ± 25.4 | - | 40.4 ± 20.0 | 53.4 ± 27.9 | 49.9 ± 32.6 | 0.27 |
| BDNF(ng/mL) | 3809 ± 1245.4 | 3596 ± 1720.5 | 3519 ± 1401.7 | 4132 ± 1337.0 | 3913 ± 1248.5 | 0.85 |
| pNF (pg/ml) | 469 ± 164.7 | 414 ± 208.7 | 365 ± 148.7 | 553 ± 163.5 | 497 ± 227.0 | 0.06 |
| IL6 (pg/ml) | 101 ± 35.7 | 122 ± 28.6 | 91 ± 48.4 | 112 ± 39.6 | 87 ± 40.4 | 0.98 |
| NT-3(pg/mL) | 11.4 ± 4.28 | 13.8 ± 3.97 | 11.0 ± 5.23 | 12 ± 7.45 | 9.63 ± 3.02 | 0.61 |
| MBP (pg/ml) | 4496 ± 1425.4 | 3950 ± 1100.5 | 3256 ± 1577.3 | 2965 ± 1032.2 | 3102 ± 1292.5 | 0.43 |
| CD95L (pg/ml) | 104 ± 52.9 | 106 ± 61.4 | 98 ± 51.4 | 107 ± 60.2 | 100 ± 62.7 | 0.99 |
| GABA (pg/ml) | 14.0 ± 5.03 | - | 14.5 ± 6.82 | 12.5 ± 5.27 | 13.1 ± 7.16 | 0.95 |
| Glutamate (pg/ml) | 24.2 ± 5.33 | - | 25.6 ± 5.78 | 27.1 ± 3.5 | 27.3 ± 6.67 | 0.24 |

*Data presented as mean ± SD or number. p-values from one-way ANOVA (continuous variables) or Fisher's exact test (categorical variables). AIS = ASIA Impairment Scale. All participants had complete motor injury (AIS grade A) with incomplete sensory sparing. No significant between-group differences were observed at baseline (all p > 0.05). LEMS = Lower Extremity Motor Score; WISCI = Walking Index for Spinal Cord Injury; SCIM = Spinal Cord Independence Measure; MT = motor threshold; CSP = cortical silent period; BDNF = brain-derived neurotrophic factor; NT-3 = neurotrophin-3; MBP = myelin basic protein; pNF = phosphorylated neurofilament heavy chain; IL-6 = interleukin-6; CD95L = CD95 ligand; BDI = Beck Depression Inventory; STAI = State–Trait Anxiety Inventory (Y1 + Y2 combined); VAS = Visual Analogue Scale for pain; WHO-QOL = WHO Quality of Life questionnaire (total score).*

## Objective 1

**Supplementary Table 2.** Effect of Stimulation Type (Placebo vs rTMS vs iTBS)

| **Variable** | **Time** |  | **Group** |  | **Time × Group** |  |
|---|---|---|---|---|---|---|
|  | **F (df)** | **P** | **F (df)** | **P** | **F (df)** | **P** |
| **Neurological** |  |  |  |  |  |  |
| LEMS | 5.36 (3,45) | **<0.01\*\*** | 0.88 (2,15) | >0.05 | 1.08 (6,45) | >0.05 |
| Light Touch | 25.46 (3,45) | **<0.001\*\*\*** | 3.20 (2,15) | >0.05 | 3.86 (6,45) | **<0.01\*\*** |
| Pin Prick | 26.80 (3,45) | **<0.001\*\*\*** | 3.43 (2,15) | >0.05 | 3.29 (6,45) | **<0.01\*\*** |
| **Functional** |  |  |  |  |  |  |
| WISCI | 20.45 (3,45) | **<0.001\*\*\*** | 6.25 (2,15) | **<0.05\*** | 6.15 (6,45) | **<0.001\*\*\*** |
| SCIM — Self-Care | 84.00 (3,45) | **<0.001\*\*\*** | 5.80 (2,15) | **<0.05\*** | 8.01 (6,45) | **<0.001\*\*\*** |
| SCIM — Resp/Sphincter | 14.41 (3,45) | **<0.001\*\*\*** | 2.17 (2,15) | >0.05 | 5.00 (6,45) | **<0.001\*\*\*** |
| SCIM — Mobility | 39.53 (3,45) | **<0.001\*\*\*** | 2.51 (2,15) | >0.05 | 5.95 (6,45) | **<0.001\*\*\*** |
| **Electrophysiological** |  |  |  |  |  |  |
| MEPmax | 5.96 (3,45) | **<0.01\*\*** | 1.96 (2,15) | >0.05 | 5.86 (6,45) | **<0.001\*\*\*** |
| I□□ (xmid) | 9.64 (3,45) | **<0.001\*\*\*** | 6.98 (2,15) | **<0.01\*\*** | 1.95 (6,45) | >0.05 |
| AUC | 15.54 (3,45) | **<0.001\*\*\*** | 6.54 (2,15) | **<0.01\*\*** | 4.89 (6,45) | **<0.001\*\*\*** |
| Cortical Silent Period | 6.65 (3,45) | **<0.001\*\*\*** | 0.42 (2,15) | >0.05 | 0.67 (6,45) | >0.05 |
| Motor Threshold | 11.16 (3,45) | **<0.001\*\*\*** | 4.27 (2,15) | **<0.05\*** | 3.51 (6,45) | **<0.01\*\*** |
| **Biochemical (3 timepoints: Pre, Post, FU)** |  |  |  |  |  |  |
| BDNF | 10.65 (2,30) | **<0.001\*\*\*** | 1.91 (2,15) | >0.05 | 4.21 (4,30) | **<0.01\*\*** |
| pNF | 6.90 (2,30) | **<0.01\*\*** | 1.48 (2,15) | >0.05 | 0.59 (4,30) | >0.05 |
| IL-6 | 28.21 (2,30) | **<0.001\*\*\*** | 0.73 (2,15) | >0.05 | 1.08 (4,30) | >0.05 |
| CD95L | 0.54 (2,30) | >0.05 | 0.04 (2,15) | >0.05 | 0.96 (4,30) | >0.05 |
| MBP | 12.53 (2,30) | **<0.001\*\*\*** | 4.20 (2,15) | **<0.05\*** | 1.50 (4,30) | >0.05 |
| NT-3 | 12.28 (2,30) | **<0.001\*\*\*** | 2.46 (2,15) | >0.05 | 1.14 (4,30) | >0.05 |
| **Psychosocial** |  |  |  |  |  |  |
| BDI | 6.98 (3,45) | **<0.001\*\*\*** | 0.22 (2,15) | >0.05 | 0.71 (6,45) | >0.05 |
| VAS | 9.76 (3,45) | **<0.001\*\*\*** | 0.16 (2,15) | >0.05 | 0.40 (6,45) | >0.05 |
| STAI-Y1 | 0.44 (3,45) | >0.05 | 3.14 (2,15) | >0.05 | 0.94 (6,45) | >0.05 |
| STAI-Y2 | 1.46 (3,45) | >0.05 | 1.10 (2,15) | >0.05 | 0.80 (6,45) | >0.05 |
| WHO-QOL Physical | 2.69 (3,45) | >0.05 | 4.72 (2,15) | **<0.05\*** | 1.80 (6,45) | >0.05 |
| WHO-QOL Psychological | 15.05 (3,45) | **<0.001\*\*\*** | 0.94 (2,15) | >0.05 | 2.51 (6,45) | **<0.05\*** |
| WHO-QOL Social | 2.73 (3,45) | >0.05 | 0.34 (2,15) | >0.05 | 1.46 (6,45) | >0.05 |
| WHO-QOL Environmental | 0.52 (3,45) | >0.05 | 0.17 (2,15) | >0.05 | 0.55 (6,45) | >0.05 |

## Objective 2

**Supplementary Table S3.** Effect of Stimulation Site (Placebo vs iTBS(MC) vs iTBS(SC) vs iTBS(MC+SC))

| Variable | Time |  | Group |  | Time × Group |  |
|---|---|---|---|---|---|---|
|  | F (df) | P | F (df) | P | F (df) | P |
| **Neurological** |  |  |  |  |  |  |
| LEMS | 8.43 (3,60) | **<0.001\*\*\*** | 2.61 (3,20) | >0.05 | 1.14 (9,60) | >0.05 |
| Light Touch | 13.93 (3,60) | **<0.001\*\*\*** | 2.48 (3,20) | >0.05 | 1.20 (9,60) | >0.05 |
| Pin Prick | 12.73 (3,60) | **<0.001\*\*\*** | 2.66 (3,20) | >0.05 | 1.28 (9,60) | >0.05 |
| **Functional** |  |  |  |  |  |  |
| WISCI | 32.60 (3,60) | **<0.001\*\*\*** | 8.37 (3,20) | **<0.001\*\*\*** | 7.41 (9,60) | **<0.001\*\*\*** |
| SCIM — Self-Care | 118.48 (3,60) | **<0.001\*\*\*** | 15.50 (3,20) | **<0.001\*\*\*** | 9.74 (9,60) | **<0.001\*\*\*** |
| SCIM — Resp/Sphincter | 24.04 (3,60) | **<0.001\*\*\*** | 2.85 (3,20) | >0.05 | 5.45 (9,60) | **<0.001\*\*\*** |
| SCIM — Mobility | 77.19 (3,60) | **<0.001\*\*\*** | 5.60 (3,20) | **<0.01\*\*** | 8.53 (9,60) | **<0.001\*\*\*** |
| **Electrophysiological** |  |  |  |  |  |  |
| MEPmax | 0.54 (3,50.9) | >0.05 | 0.99 (3,20.1) | >0.05 | 2.67 (9,50.6) | **<0.05\*** |
| I□□ (xmid) | 6.27 (3,51.4) | **<0.01\*\*** | 5.46 (3,19.2) | **<0.05\*** | 0.86 (9,50.9) | >0.05 |
| AUC | 21.73 (3,60) | **<0.001\*\*\*** | 7.10 (3,20) | **<0.01\*\*** | 5.30 (9,60) | **<0.001\*\*\*** |
| Motor Threshold | 15.76 (3,60) | **<0.001\*\*\*** | 3.20 (3,20) | **<0.05\*** | 2.79 (9,60) | **<0.01\*\*** |
| Cortical Silent Period | 16.46 (3,60) | **<0.001\*\*\*** | 0.61 (3,20) | >0.05 | 1.58 (9,60) | >0.05 |
| LICI | 9.51 (3,924) | **<0.001\*\*\*** | 2.38 (3,20) | >0.05 | 5.64 (9,924) | **<0.001\*\*\*** |
| SICI-ICF (combined) | 10.88 (3,924) | **<0.001\*\*\*** | 11.86 (3,20) | **<0.001\*\*\*** | 4.20 (9,924) | **<0.001\*\*\*** |
| **Biochemical (3 timepoints: Pre, Post, FU)** |  |  |  |  |  |  |
| pNF | 9.09 (2,40) | **<0.001\*\*\*** | 1.22 (3,20) | >0.05 | 4.22 (6,40) | **<0.01\*\*** |
| MBP | 7.66 (2,40) | **<0.01\*\*** | 2.61 (3,20) | >0.05 | 1.03 (6,40) | >0.05 |
| BDNF | 1.38 (2,40) | >0.05 | 2.65 (3,20) | >0.05 | 3.19 (6,40) | **<0.05\*** |
| NT-3 | 12.10 (2,40) | **<0.001\*\*\*** | 1.44 (3,20) | >0.05 | 4.31 (6,40) | **<0.01\*\*** |
| IL-6 | 29.40 (2,40) | **<0.001\*\*\*** | 0.90 (3,20) | >0.05 | 0.79 (6,40) | >0.05 |
| CD95L | 0.98 (2,40) | >0.05 | 0.04 (3,20) | >0.05 | 0.75 (6,40) | >0.05 |
| **Psychosocial** |  |  |  |  |  |  |
| WHO-QOL Physical | 4.73 (3,60) | **<0.01\*\*** | 4.35 (3,20) | **<0.05\*** | 1.52 (9,60) | >0.05 |
| WHO-QOL Psychological | 41.90 (3,60) | **<0.001\*\*\*** | 2.27 (3,20) | >0.05 | 5.76 (9,60) | **<0.001\*\*\*** |
| WHO-QOL Social | 3.43 (3,60) | **<0.05\*** | 0.43 (3,20) | >0.05 | 1.48 (9,60) | >0.05 |
| WHO-QOL Environmental | 3.97 (3,60) | **<0.05\*** | 0.58 (3,20) | >0.05 | 1.72 (9,60) | >0.05 |
| BDI | 12.27 (3,60) | **<0.001\*\*\*** | 0.36 (3,20) | >0.05 | 1.22 (9,60) | >0.05 |
| VAS | 16.05 (3,60) | **<0.001\*\*\*** | 0.17 (3,20) | >0.05 | 0.64 (9,60) | >0.05 |
| STAI-Y1 | 0.70 (3,60) | >0.05 | 2.69 (3,20) | >0.05 | 0.37 (9,60) | >0.05 |
| STAI-Y2 | 0.75 (3,60) | >0.05 | 0.25 (3,20) | >0.05 | 1.02 (9,60) | >0.05 |

*\*P<0.05, \*\*P<0.01, \*\*\*P<0.001. Type III Analysis of Variance with Satterthwaite's method. F values followed by degrees of freedom in parentheses (numerator df, denominator df). Biochemical outcomes assessed at 3 time points (Pre, Post, FU) only. LICI, SICI, and ICF models were fit on the full repeated interstimulus-interval (ISI) dataset (924). LICI = long-interval intracortical inhibition; SICI = short-interval intracortical inhibition; ICF = intracortical facilitation; GABA = gamma-aminobutyric acid; LEMS = Lower Extremity Motor Score; WISCI = Walking Index for Spinal Cord Injury; SCIM = Spinal Cord Independence Measure; AUC = area under the curve; WHO-QOL = World Health Organization Quality of Life; BDI = Beck Depression Inventory; VAS = Visual Analogue Scale; STAI = State-Trait Anxiety Inventory.*

