## Supplementary method for "Reactivating Residual Motor Circuits in Complete Spinal Cord Injury Through Mode- and Site-Specific Transcranial Magnetic Stimulation: A Pilot Randomized Controlled Trial"

**Supplementary Methods S1. Stimulation Protocols**

#### S1.1 Coil Positioning and Motor Hotspot Localisation for TMS intervention

The primary motor cortex leg representation was localised using the international 10–20 EEG system as a reference. The centre of the figure-of-eight coil was positioned at the vertex (Cz electrode site), corresponding to the approximate location of the leg primary motor cortex. The Placebo, rTMS and iTBS stimulation was delivered at leg primary motor cortex. The optimal coil position was marked on a close-fitting cap worn by the participant to ensure consistent placement across sessions. In iTBS-MC group the coil targeted the leg area of the M1; iTBS-SC group it targeted the thoracolumbar region at injury level; and in iTBS-MC+SC group, the coil was applied at both sites sequentially with a 2-minute interval. rTMS was delivered at 20 Hz (2-second trains, 28-second inter-train interval (ITI)), 1600 pulses/session; total time duration of 20min) and iTBS was delivered at 600 pulses/session (50 Hz bursts at 5 Hz, 2-second trains, 8-second ITI; total duration 3.20 min).

For spinal cord stimulation (iTBS-SC) and the spinal component of iTBS-MC+SC group, the coil was positioned over thoracolumbar spine, centred at the vertebral level, targeting the injury site. Injured SC level was determined by the CT report and clinical neurological level of injury. The coil was held parallel to the spinal axis, targeting dorsal column afferents and segmental interneuronal circuits. Coil-to-skin distance was minimised and maintained consistently across sessions using the same positioning landmarks.

#### S1.2 Placebo Stimulation Procedure

The Placebo stimulation was delivered using the active figure-of-eight coil, placed on the hotspot at 90° to the scalp. The coil was connected with the TMS machine module which has a inbuilt software (Neuro-MS/MD) to give placebo stimulation, thereby preventing any cortical activation while replicating the auditory click and cutaneous sensations associated with active TMS. The stimulation parameters were similar to iTBS (frequency, intensity, train duration, inter-train interval).

#### S1.3 Resting Motor Threshold Determination

Resting motor threshold (RMT) was determined prior to each electrophysiological assessment. Surface EMG electrodes were placed over the APB muscle belly (active electrode) and the proximal interphalangeal joint of the first digit (reference electrode). The participants were instructed to maintain complete muscle relaxation, verified by live EMG monitoring. Single TMS pulses were delivered in a step-wise fashion starting at 20% maximum stimulator output (MSO) and incrementing by 5% until MEPs were evoked. RMT was defined as the minimum intensity capable of evoking MEPs with a peak-to-peak amplitude of ≥ 50 µV in at least 5 of 10 consecutive trials.

#### S1.4 Session Structure and Safety Monitoring

Each stimulation session lasted approximately 20 minutes (rTMS) or 3 minutes (iTBS). Electrode placement, recording of motor threshold, and other electrophysiological parameters took additional 30-40 minutes. Sessions were delivered twice daily with a minimum inter-session interval of two hours. For the combined condition (iTBS-MC+SC), motor cortex stimulation was always delivered first, followed by spinal cord stimulation within the same session. Participants were seated comfortably in a supported chair during cortical stimulation and in a prone or supported lying position during spinal stimulation.

Safety was monitored throughout the study. Vital signs (blood pressure, heart rate, oxygen saturation) were recorded before and after each session. Participants were screened for adverse events at each session using a standardised TMS safety checklist including headache, nausea, neck pain, syncope, muscle twitching, seizure-like activity, skin irritation at electrode or coil contact sites, and any other discomfort. 2 patients complaint for mild headache after intervention, that vanish after 15 minutes of intervention. No TMS-related seizures or serious adverse events occurred during the study.

#### S1.5 Rehabilitation Protocol

All participants received a standardised rehabilitation programme of 45 minutes per session, delivered after each stimulation session by a qualified physiotherapist blinded to group allocation. The rehabilitation programme comprised of:

**Upper limb strengthening (15 minutes):** Resisted exercises targeting shoulder, elbow, and wrist using theraband and free weights, progressed according to participant tolerance.

**Lower limb mobilisation (15 minutes):** Passive range-of-motion exercises for hip, knee, and ankle joints; active-assisted movements where any voluntary activity was present; stretching for hip flexors and ankle plantar flexors.

**Bed mobility and transfer training (5 minutes):** Rolling, supine-to-sit transitions, and wheelchair transfer techniques.

**Functional task training (10 minutes):** Task-oriented activities targeting activities of daily living, including reaching, grasping, and supported stepping where applicable.

No additional physiotherapy, TMS, or other neurostimulation was permitted during the intervention period.

### Supplementary Methods S2. Neurological and Functional Parameters

### S2.1 ASIA Impairment Scale (AIS scale)

### The motor examination assesses ten key muscle groups, representing major cervical and lumbosacral myotomes. These include five upper-limb muscles—elbow flexors (C5), wrist extensors (C6), elbow extensors (C7), finger flexors (C8), and little finger abductors (T1)—and five lower-limb muscles—hip flexors (L2), knee extensors (L3), ankle dorsiflexors (L4), long toe extensors (L5), and ankle plantar flexors (S1). Muscle strength is graded bilaterally on a 6-point scale ranging from 0 (total paralysis) to 5 (normal strength). The maximum motor score is 100, with 50 points assigned to each side of the body.

### The sensory examination evaluates 28 dermatomes on both sides of the body for light touch (using cotton) and pinprick sensation (using a sterile safety pin). Sensation is scored as 0 (absent), 1 (impaired), or 2 (normal). A normal sensory examination yields a maximum score of 224, comprising 112 points from each side. Inability to distinguish pinprick from light touch is scored as absent sensation (0).

### As the participants with spinal cord injury in the present study were paraplegic, motor function was assessed using only the Lower Extremity Motor Score (LEMS).

### S2.2 Walking index for SCI (WISCI)-II

### Walking ability was assessed using the Walking Index for Spinal Cord Injury-II (WISCI-II), a 21-level ordinal scale (0–20) that measures the amount of physical assistance, braces, and assistive devices required to walk 10 m. Higher scores reflect greater walking independence and functional recovery.

### S3.3 Spinal cord independence measure -III (SCIM)

### Functional independence was assessed using the Spinal Cord Independence Measure. SCIM is a spinal cord injury-specific questionnaire designed to evaluate activities of daily living and functional independence. The SCIM-III consists of three domains: self-care (0–20 points), respiration and sphincter management (RSM; 0–40 points), and mobility (0–40 points), yielding a total score ranging from 0 to 100. The self-care domain assesses feeding, grooming, bathing, and dressing activities. The RSM domain evaluates respiratory function, bladder management, bowel management, and toilet use. The mobility domain assesses transfers, bed mobility, and indoor and outdoor locomotion. Higher scores in each domain indicate greater functional independence.

### Supplementary Methods S3. Detailed Electrophysiological Parameters Recording

#### S3.1 EMG Recording Setup

Surface EMG was recorded using disposable Ag/AgCl electrodes (10 mm diameter) placed in a belly of the APB muscle. The ground electrode was placed over the dorsum of the hand. EMG signals were amplified (× 1000), bandpass filtered (20–2000 Hz), and digitised at 5 kHz using a 16-bit analogue-to-digital converter. Data were stored offline for analysis.

All the electrophysiological parameters recording were taken from the APB muscle. To localise the motor hotspot for APB, figure-of-eight coil was placed at vertex using the international 10–20 EEG system. Fine adjustment was performed by moving the coil in 5 cm lateral axis and 2 cm anterior axis to identify the motor area for abductor pollicis brevis (APB) muscle. To identify the site of maximum corticospinal excitability point in motor cortex, 1cm anterio-posterior and mediolateral adjustment was done while recording RMT from APB muscle. Coil orientation was maintained with the handle pointing posterolateral at approximately 45° to the midsagittal plane, inducing a posterior-to-anterior current flow in the underlying cortex.

#### S3.2 Resting Motor Threshold

RMT was defined as the minimum stimulus intensity capable of evoking MEPs with a peak-to-peak amplitude exceeding 50 µV in at least 5 of 10 consecutive trials. RMT was used as the conditioning stimulus intensity for SICI and ICF protocols.

#### S3.3 MEP Recruitment Curves

MEPs were recorded from APB using surface EMG (20–2000 Hz bandpass; 5 kHz sampling). Recruitment curves (RC) were generated at 10–100% MSO (10% steps; 5 stimuli/intensity). MEP amplitudes were fitted to a four-parameter Boltzmann sigmoidal function:

**MEP(x) = MEP**_max_ **/ {1 + exp[−(x − I**_50_**) / k]}**

where x = stimulus intensity (%MSO), MEP_max_ = maximum MEP amplitude (mV), I_50_ = intensity at 50% MEP_max_ (%MSO), and k = slope parameter. Fitting used non-linear least-squares model (Levenberg–Marquardt; R^2^ ≥ 0.80 required) and AUC was computed by trapezoidal integration. RMT was defined as the minimum intensity evoking MEPs ≥ 50 µV in ≥ 5/10 trials. CSP duration was measured at 120% RMT during voluntary APB contraction. Paired-pulse TMS parameters (SICI: ISI 1–5 ms; ICF: ISI 10–30 ms; LICI: ISI 100–300 ms) were assessed only in Objective 2 following equipment availability.

#### S3.4 Cortical Silent Period

CSP was assessed during voluntary contraction of the APB at approximately 30% of maximum voluntary contraction (MVC). A single suprathreshold TMS pulse (120% RMT) was delivered, and 10 trials were recorded. The cortical silent period (cSP, msec) was calculated from the onset of EMG suppression or silencing till reoccurrence of MEP during voluntary muscle contraction.

**S3.5 Paired-Pulse TMS parameters**

***Short-Interval Intracortical Inhibition (SICI) and Intracortical Facilitation (ICF)***

SICI and ICF were assessed using a paired-pulse TMS paradigm delivered through the Neuro-MS/MD system. A subthreshold conditioning stimulus (CS) set at 80% of resting motor threshold (RMT) preceded a suprathreshold test stimulus (TS). Rather than fixing the TS at RMT, the TS intensity was individually calibrated to the stimulator output required to produce a peak-to-peak MEP amplitude of 1 mV in the target muscle — an approach that reduces inter-participant variability in assessing corticospinal excitability more reliably than RMT-based targeting. This 1 mV MEP served as the individual reference (100%), against which all conditioned responses were expressed.

Interstimulus intervals (ISIs) of 1, 2, 3, 4, and 5 ms were used for SICI, and ISIs of 10, 15, 20, and 30 ms were used for ICF. Ten trials were recorded at each ISI. Conditioned MEP amplitudes were expressed as a percentage of the unconditioned 1 mV reference.

**%MEP = {Conditioned MEP amplitude/ Unconditioned (1 mV) MEP amplitude} × 100**

- 100% MEP → no inhibition; conditioned MEP equals the test MEP.
- <100% MEP → inhibition; the conditioning pulse suppressed the corticospinal response.

***Lower the percentage**, **stronger the inhibition**.

***Long-Interval Intracortical Inhibition (LICI)***

LICI was assessed using two suprathreshold stimuli, both at 120% RMT, with ISIs of 100, 150, 200, and 300 ms. Ten trials per ISI were recorded. Mean LICI was calculated across all four ISIs. All paired-pulse assessments were conducted by the same operator, and a minimum of 5 minutes separated the SICI/ICF and LICI blocks to prevent carry-over effects.

### Supplementary Methods S4. Biochemical Assays

**S4.1 Enzyme linked immunosorbent assay (ELISA )**

Peripheral venous blood samples were collected from all participants at each pre-intervention, post intervention and 2^nd^ follow-up. After collection, the samples were allowed to clot at room temperature for 30 minutes and then centrifuged at 3,000 rpm for 10 minutes to separate the serum. The serum was aliquoted and stored at −80°C until analysis. Before testing, samples were thawed on ice and used after a single freeze–thaw cycle.

Serum levels of brain-derived neurotrophic factor (BDNF), neurotrophin-3 (NT-3), phosphorylated neurofilament heavy chain (pNF-H), myelin basic protein (MBP), interleukin-6 (IL-6), and CD95 ligand (CD95L) were measured using commercially available ELISA kits (ImmunoTag™, GBiosciences, USA) according to the manufacturer's instructions.

For each assay, standards and serum samples were added to antibody-coated microplate wells and incubated to allow binding of target analyte. After washing, enzyme-linked detection antibodies were added, followed by substrate solution for colour development. The reaction was stopped using stop solution, and absorbance was measured at 450 nm using a microplate reader. Biomarker concentrations were determined from standard curves generated for each plate using a four-parameter logistic (4PL) model. All samples were analysed in duplicate, and the mean value was used for statistical analysis.

**S4.2 Liquid chromatography–tandem mass spectrometry (LC-MS/MS)**

Plasma GABA and glutamate were quantified by LC-MS/MS using a triple quadrupole mass spectrometer coupled to HPLC. EDTA plasma samples were deproteinised with cold acetonitrile (1:3 v/v), centrifuged at 14,000 × g for 10 min at 4°C, and the supernatant derivatised immediately with OPA/MPA at room temperature for 2 minutes. Separation was performed on a reverse-phase C18 column (150 × 2.1 mm, 3.5 µm) using a gradient of 10 mM ammonium formate pH 3.5 (A) and acetonitrile/methanol 90:10 (B): 5% B for 0–2 min, ramped to 95% B over 2–8 min, held to 10 min, then re-equilibrated. MRM transitions were 104→87 m/z (quantifier) and 104→69 m/z (qualifier) for GABA, and 148→130 m/z and 148→84 m/z for glutamate. Isotope-labelled internal standards (GABA-d6 and glutamate-d5) were used to correct for matrix effects, with calibration ranges of 0.1–100 µmol/L and 1–500 µmol/L respectively. All samples were analysed in a single batch to minimise inter-run variability.

### Supplementary Methods S5. Psychosocial Parameters

### World Health Organization Quality of life-Brief (WHOQOL-BREF)

### Quality of life was assessed using the WHOQOL-BREF questionnaire, which consists of four domains: Physical Health (7 items), Psychological Health (6 items), Social Relationships (3 items), and Environment (8 items). Each item is rated on a 5-point Likert scale (1–5), with higher scores indicating better quality of life. The physical domain evaluates mobility, activities of daily living, and sleep; the psychological domain assesses emotional well-being, self-esteem, and cognitive functioning; the social domain evaluates personal relationships and social support; and the environmental domain assesses safety, financial resources, healthcare accessibility, opportunities for learning and leisure, and the physical environment. Domain scores were calculated according to WHOQOL-BREF guidelines, with higher scores reflecting better quality of life.

### Beck depression inventory-II (BDI)

### Beck Depression Inventory-II (BDI-II) is a 21-item self-report questionnaire widely used to evaluate the severity of depression. Each item is scored on a 4-point scale ranging from 0 to 3, with total scores ranging from 0 to 63. Higher scores indicate greater severity of depressive symptoms. Total scores are categorized as minimal (0–13), mild (14–19), moderate (20–28), and severe depression (29–63).

### Visual Analogue Scale (VAS)

### Pain intensity was assessed using the Visual Analogue Scale (VAS). The VAS consists of a 10-cm horizontal line anchored by “no pain” at one end and “worst imaginable pain” at the other. Participants were asked to mark the point on the line that best represented their current pain intensity. Scores ranged from 0 to 10, with higher scores indicating greater pain severity.

### The Spielberger State-Trait Anxiety Inventory (STAI)

### Anxiety was assessed using the Spielberger State–Trait Anxiety Inventory (STAI), a self-report questionnaire comprising two 20-item subscales: State Anxiety form Y-1 (STAI-S) and Trait Anxiety form Y-2 (STAI-T). The State Anxiety scale measures the individual's current level of anxiety (how they feel right now), whereas the Trait Anxiety scale assesses the general tendency to experience anxiety (how they generally feel). Each item is rated on a 4-point Likert scale, with total scores ranging from 20 to 80 for each subscale. Higher scores indicate greater levels of anxiety.
