## supplementary results for "Reactivating Residual Motor Circuits in Complete Spinal Cord Injury Through Mode- and Site-Specific Transcranial Magnetic Stimulation: A Pilot Randomized Controlled Trial"

**Objective 1: Effect of Stimulation Mode**

### Neurological outcomes

### In sensory function, a significant time X group interactions was observed for both the interventions (rTMS-FU1: p = 0.0007; FU2: p = 0.0003 and iTBS -FU2: p = 0.0196). Post-hoc analysis showed a significant increase in scores of iTBS group at FU2 in light touch (p = 0.047) and pin-prick (p = 0.039) sensation as compared to placebo.

Temporal within group comparisons revealed significant improvements in sensory scores of light touch in rTMS (FU1: p = 0.0007; FU2: p = 0.0003) and iTBS groups (FU1: p = 0.0196; FU2: p = 0.0045) as compared to baseline. The pin prick score also showed a significant increase following rTMS (FU1: p = 0.0035; FU2: p = 0.0007) and iTBS (FU1: p = 0.0331; FU2: p = 0.0043) intervention.

**b**

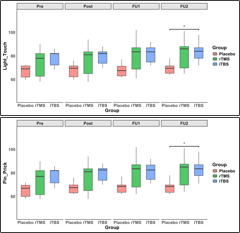

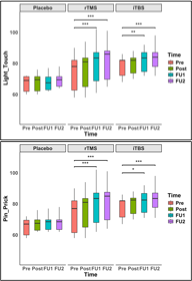

**a**

### Supplementary Figure_S1. AIS Sensory Scores (median with interquartile range) recovery was evaluated using 2 domains of AIS: Light touch and Pin prick, across four time points: pre-intervention (Pre), post-intervention (Post), first follow-up (FU1), and second follow-up (FU2). (a) between the group comparisons of LEMS at each time point. (b) temporal within group comparisons. *p < 0.05, **p < 0.01 and ***p < 0.001.

### Functional outcomes: SCIM domain scores

**Self-Care:** Significant within group improvements were found in iTBS (Post: p = 0.004; FU1:p < 0.001; FU2: p < 0.001) as well as in rTMS groups (Post: p = 0.03; FU1: p = 0.002; FU2: p < 0.001). Placebo group also improved significantly from baseline to FU2 (p = 0.007). A significant beneficial effect of both rTMS and iTBS interventions was evident at FU1 and FU2 (all p < 0.05) as compared to placebo.

**Respiration and Sphincter Management:** Within-group analysis showed significant improvements in both rTMS (FU2: p = 0.002) and iTBS groups (FU1: p = 0.003; FU2: p = < 0.001). Between-group analysis showed increase scores in iTBS (FU: p = 0.029; FU2: p = 0.001) compared to placebo.

**Mobility:** Both rTMS (FU1: p = 0.01 and FU2: p = 0.007 ) and iTBS (FU1: p = 0.013 and FU2: p < 0.001) showed a significant within group improvement. At FU2, between group analysis confirmed the significant increase in mobility score in iTBS compared to placebo (p < 0.001) and rTMS (p = 0.049).

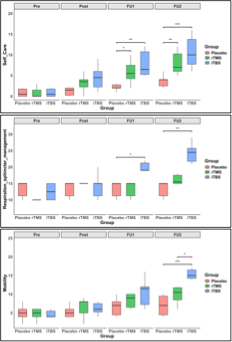

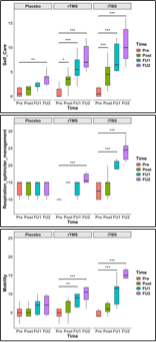

**a**

**b**

### Supplementary Figure_S2. Spinal Cord Independence Measure (SCIM, median with interquartile range) was assessed in 3 domains (Selfcare, Respiration sphincter management and mobility) across four time points: pre-intervention (Pre), post-intervention (Post), first follow-up (FU1), and second follow-up (FU2). (a) intergroup comparisons at each time point. (b) within-group temporal changes in SCIM scores. Statistical annotations: *p < 0.05, **p < 0.01, ***p < 0.001.

**Electrophysiological outcomes**

**Recruitment curve parameters:**

**Area under the curve (AUC)-** AUC was significantly higher in iTBS group (Post: p = 0.004; FU1: p < 0.001; FU2: p = 0.020), and in rTMS (Post: 0.046; FU1: p = 0.006) as compared to placebo. Within-group analysis showed a significant increase in AUC for both rTMS (FU1: p = 0.019) and iTBS (Post: p = 0.018; FU1: p < 0.001; FU2: p = 0.020) groups.

**MEPmax-** Increased MEPmax observed in iTBS group as compared to placebo at FU1 (p = 0.008). Within-group analysis revealed significant increase in MEP_max_ in both rTMS (Post: p = 0.027) and iTBS (Post: p = 0.013; FU1: p < 0.001; FU2: p = 0.0003) groups over the observation period.

**I₅₀ (Stimulus Intensity at 50% MEPmax)-** I₅₀ decreased significantly following iTBS intervention as compared to placebo at post (p = 0.0005) and FU1 (p = 0.022), indicating less intensity required to elicit 50% MEP amplitude. Within group analysis showed a significant reduction in I₅₀ in rTMS (Post: p = 0.006; FU1: p = 0.0002; FU2: p = 0.003), and iTBS groups (Post: p = 0.027; FU1: p = 0.024) over the time points.

**a**

**b**

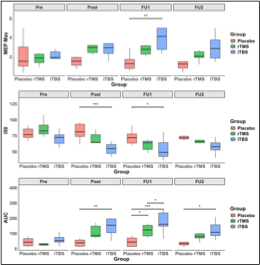

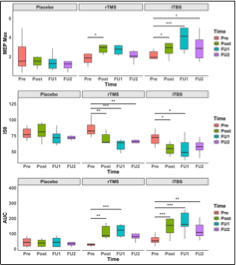

**Supplementary Figure_S3. MEP recruitment curve parameters** (median with interquartile range, MEPmax (mV), I50 (%), and area under the curve (AUC) across four time points: pre-intervention (Pre), post-intervention (Post), first follow-up (FU1), and second follow-up (FU2). **(a)** inter**group comparisons** at each time point. **(b)** **within-group comparisons** across time points. Statistical annotations: *p < 0.05, **p < 0.01, ***p < 0.001.

**Cortical Silent Period:** Within-group analysis showed reduction in cSP in both iTBS (Post: p = 0.030; FU1: p = 0.012) and rTMS (Post: p = 0.022), indicating modulation of intracortical inhibitory activity in both active groups. No significant between-group difference at any time point.

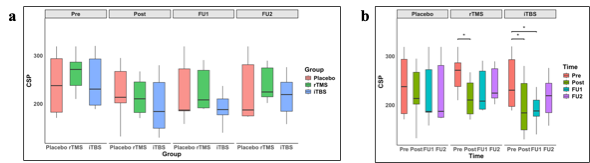

### Supplementary Figure_S4. Cortical silent period (msec; Median with interquartile range) across four time points: pre-intervention (Pre), post-intervention (Post), first follow-up (FU1), and second follow-up (FU2). (a) intergroup comparisons at each time points (b) t within-group comparison over the time points. *p < 0.05.

### Serum biomarkers

### Between group comparison revealed a significant decrease in MBP at post intervention and FU (p < 0.05); a significant increase in BDNF (p = 0.004) and NT-3 (p = 0.036) levels in iTBS groups in comparison with placebo at FU.

### Within group temporal comparisons revealed, in iTBS group a significant decrease in pNF (Pre-Post: p = 0.011), MBP (Pre–Post: p = 0.005; Pre–FU: p = 0.002) and IL-6 (Pre–Post: p = 0.038; Pre–FU: p < 0.001), whereas, a significant increase was seen in the levels of BDNF (Pre–FU: p = 0.0001) and NT-3 (Pre–Post: p = 0.018; Pre–FU: p = 0.0003) as compared to baseline.

rTMS group also a significant decrease in MBP (Pre–FU: p = 0.006) and IL-6 (Pre–FU: p = 0.016) levels was observed, whereas, the levels increased significantly of BDNF (Pre–Post: p = 0.031; Pre–FU: p = 0.011) and NT-3 (Pre–FU: p = 0.050) as compared to baseline.

A significant within-group decrease in IL-6 levels were also observed in placebo (Pre–FU: p = 0.001), indicating a generalised time-related anti-inflammatory response across all groups.

No significant within-group or between-group differences were observed in CD95L levels.

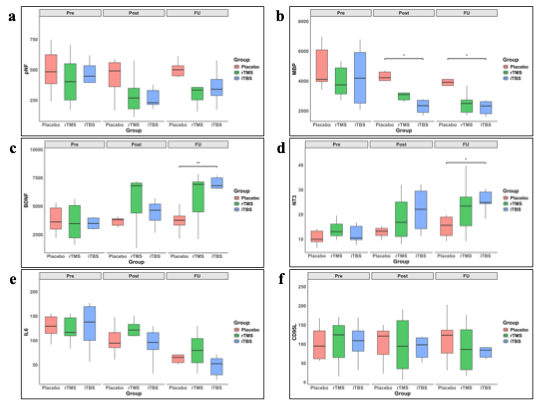

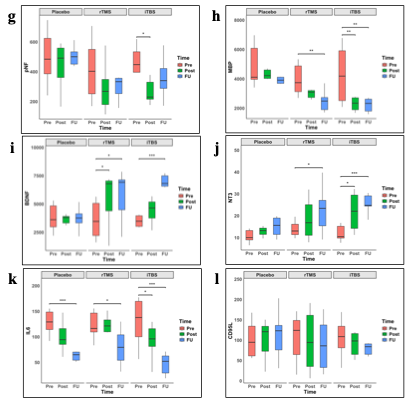

**Supplementary Figure_S5. Levels of various serum biomarkers (**pg/ml, Median with interquartile range**) (a–f)** present comparisons among the groups across various time points; and **(g–l)** illustrate within-group temporal changes across the same time points for each intervention. (c, i) BDNF, (d, j) NT3, (e, k) IL-6, and (f, l) CD95L. Data are presented as boxplots (median, and interquartile range). *p ≤ 0.05, **p ≤ 0.01, ***p ≤ 0.001.

### Psychosocial outcomes

WHO-Quality of Life (WHOQOL) domains

*Physical Domain:* At FU2, both rTMS (p = 0.017) and iTBS (p = 0.007) were significantly higher than placebo. A significant within-group improvement was also observed in iTBS (Pre–FU2: p = 0.035). *Psychological Domain:* iTBS group showed significantly higher score than placebo (FU2: p = 0.045). Within group temporal comparison revealed a significant increase in the score in both rTMS (Pre–Post: p = 0.015; Pre–FU1: p = 0.002; Pre–FU2: p < 0.001) and iTBS groups (Pre–FU1: p = 0.009; Pre–FU2: p = 0.0002) in comparison to baseline.

*Social Domain:* A significant within-group improvement was observed in iTBS group only (Pre–FU2: p = 0.023). *Environmental Domain:* No significant within-group or between-group differences were observed in any group.

*Beck Depression Inventory (BDI):* Significant reductions in BDI scores were observed in rTMS (Pre–FU2: p = 0.033) and iTBS (Pre–FU1: p = 0.033; Pre–FU2: p = 0.004).

*Visual Analogue Scale for Pain (VAS)* also a significant decrease was observed in rTMS (Pre–FU2: p = 0.037) and iTBS groups (Pre–FU1: p = 0.013; Pre–FU2: p = 0.001).

However, no changes were evident in *State Anxiety (STAI-Y1 and Y2)* scores.

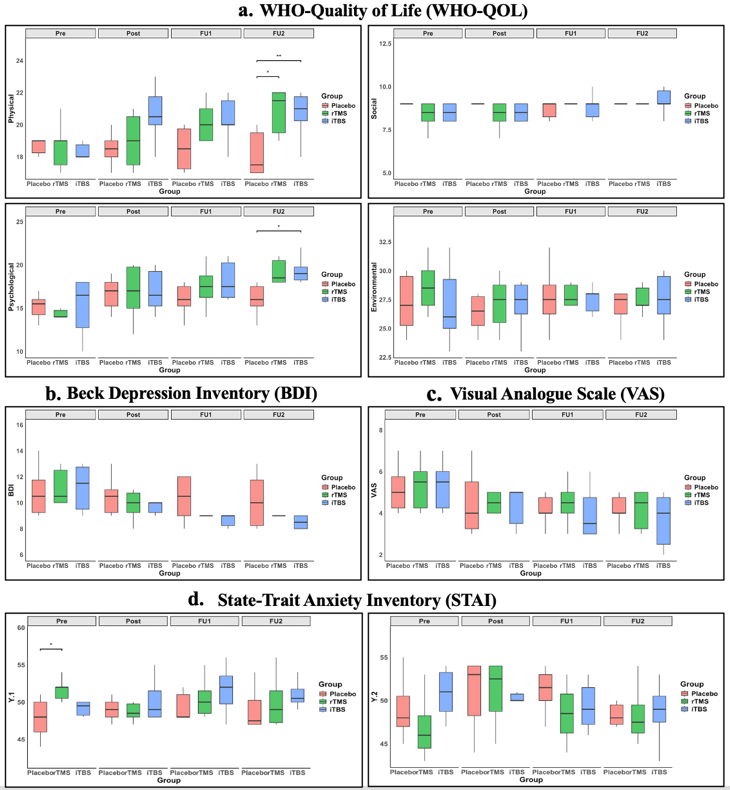

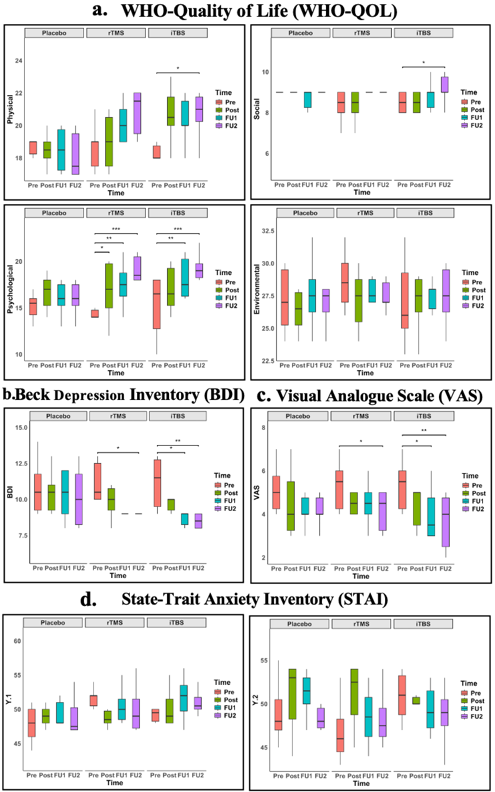

**Supplementary Figure_S6.** **Psychosocial outcomes across four time points** (Median with interquartile range): pre-intervention (Pre), post-intervention (Post), first follow-up (FU1), and second follow-up (FU2), for each group (Placebo, rTMS, iTBS). (a-d) comparisons among the groups; and (e-h) temporal within-group comparisons. *p < 0.05, **p < 0.01, ***p < 0.001.

**Correlation Analysis:**

ΔLEMS showed a significant positive correlation with ΔWISCI (R = 0.68, p = 0.002). mobility subdomain of SCIM (R = 0.66, p = 0.003) positively correlated with ΔWISCI. ΔWISCI is also significantly corelated with the physical subdomain of WHOQOL (R = 0.65, p = 0.003). ΔWISCI was positively correlated with ΔMEP (R = 0.62, p = 0.006), and ΔAUC (R = 0.61, p = 0.007). Positive correlation was observed in ΔNT3 and ΔWISCI (R = 0.53, p = 0.023).

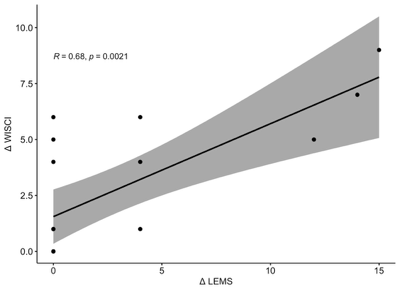

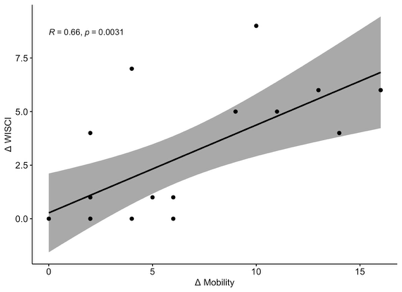

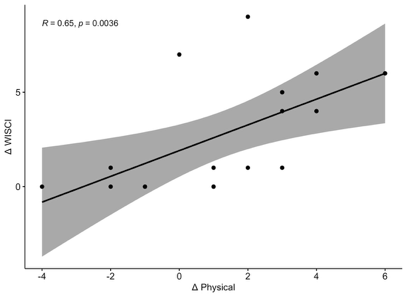

a.

c.

b.

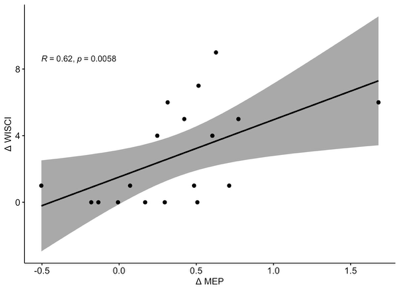

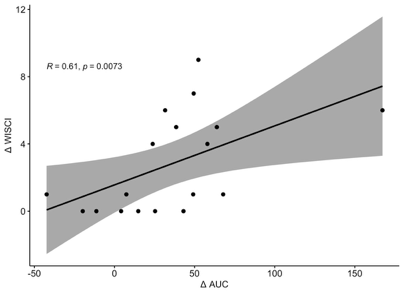

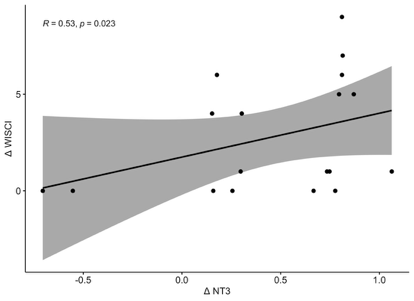

f.

e.

d.

**Supplementary Figure_S7. Spearman correlation analysis between change scores (Δ) in neurological, functional, electrophysiological, and biochemical outcomes following intervention.**

ΔWISCI correlated positively with (a) ΔLEMS, (b) ΔMobility, (c) ΔWHO-QOL Physical, (d) ΔMEP amplitude, (e) ΔAUC, and (f) ΔNT-3. Shaded bands represent 95% confidence intervals. R = Spearman's rank correlation coefficient, p = p-value, indicating the statistical significance of the correlation. LEMS = Lower Extremity Motor Score; WISCI = Walking Index for Spinal Cord Injury; WHO-QOL = World Health Organization Quality of Life; MEP = motor evoked potential; AUC = area under the curve; NT-3 = neurotrophin-3.

**Objective 2: Effect of iTBS stimulation site on outcome parameters**

### Neurological outcomes

**Light Touch:** Between-group analysis showed higher light touch score in iTBS(MC) groups as compared to placebo at FU2 (p = 0.048). Temporal within group comparison observed a significant improvement at FU2 in iTBS (MC) (p = 0.007), iTBS (SC) (p = 0.009) and iTBS (MC+SC) (p = 0.009) groups as compared to their baseline values. iTBS (MC+SC) group also showed significant improvement at FU1 with respect to baseline and post-intervention (Pre–FU1: p = 0.007).

**Pin Prick**: A significant increase in pin prick scores were observed in iTBS (MC) as compared to placebo at FU2 (p = 0.044). In comparison to baseline, a significant improvement was found in iTBS (MC) (Pre–FU2: p = 0.028), iTBS(SC) (Pre–FU2: p = 0.008), and iTBS (MC+SC) (Pre–FU1: p = 0.0003; Pre–FU2: p = 0.003) groups.

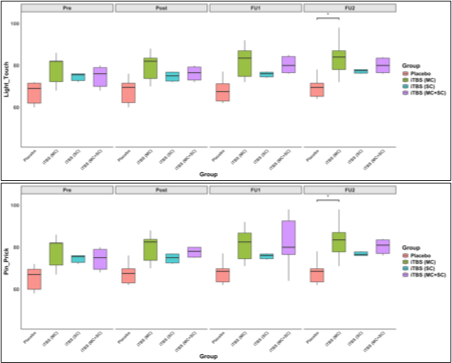

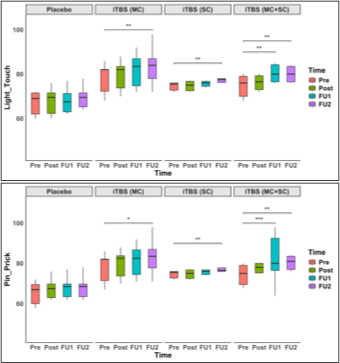

**a**

**b**

### Supplementary Figure_S8. AIS Sensory Score (median with interquartile range) was evaluated using 2 domains of AIS: Light touch and Pin prick, across four time points: pre-intervention (Pre), post-intervention (Post), first follow-up (FU1), and second follow-up (FU2). (a) represent between-group comparisons across Placebo, iTBS (MC), iTBS (MC+SC), and iTBS (SC) groups at each time point. (b) depict within-group changes. *p < 0.05, **p < 0.01 and ***p < 0.001.

### Functional outcomes: SCIM domain scores

**Self-Care**: All three active iTBS groups showed significant improvements at all-time points as compared to placebo (p < 0.05). iTBS(MC+SC) significantly increased as compared to iTBS (SC) at Post, FU1 and FU2 (p < 0.05). All groups also showed a significant improvement as compared to baseline (p <0.05).

**Respiration and Sphincter Management:** iTBS(MC) and iTBS(MC+SC) groups had higher scores in comparison to placebo at FU2 (p = 0.0007, p < 0.0001). Further, significant improvements (p < 0.01) were also observed in iTBS(MC) and iTBS(MC+SC) groups at FU1 and FU2 as compared to baseline.

**Mobility scores** significantly increased in iTBS(MC) and iTBS(MC+SC) groups as compared to placebo at FU2 (p = 0.0001 and p < 0.0001). In comparison to baseline data, the mobility scores were found to be significantly higher in iTBS(MC+SC) (Post: p = 0.012; FU1: p < 0.001; FU2: p < 0.001); iTBS(MC) (FU1: p = 0.021; FU2: p < 0.001), iTBS(SC) (FU2: p = 0.009) groups.

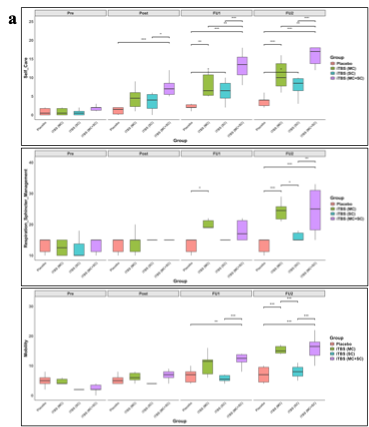

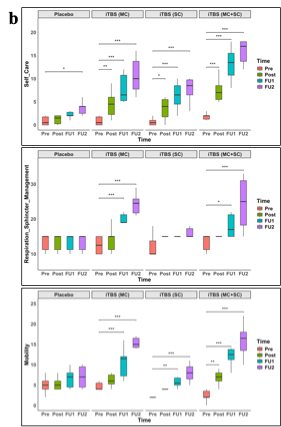

### Supplementary Figure_S9. Spinal cord independence measure (SCIM) domains (Selfcare, Respiration sphincter management and mobility) was assessed at four time points (a) comparison among the group at each time point. (b) within-group longitudinal changes in SCIM scores across time. *p < 0.05, **p < 0.01, ***p < 0.001.

**Electrophysiological outcomes**

**Recruitment curve parameters:**

*Area under the curve (AUC):* AUC was significantly higher in iTBS (MC) and iTBS(MC+SC) compared to placebo at Post (p = 0.0027; p = 0.0024), FU1 (p < 0.001; p = 0.016), and FU2 (p = 0.018; p = 0.021). AUC of iTBS (MC) and iTBS(MC+SC) was also higher than iTBS (SC) at post (p = 0.0132; p = 0.011) and at FU1 (iTBS (MC) vs iTBS (SC) p = 0.0002; p = 0.011). Within group comparisons also revealed higher (p < 0.01) AUC at all time points in iTBS (MC) and iTBS(MC+SC) in comparison to baseline.

*MEP_max_:* iTBS(MC) increased significantly as compared to placebo at FU1 (p = 0.035). In comparison to baseline, MEP_max_ increased significant in iTBS(MC) at FU1 (p = 0.015), and placebo (FU1: p = 0.022; FU2: p = 0.016) .

*I₅₀:* I₅₀ decreased significantly in iTBS(MC) compared to placebo at Post (p = 0.0009). within group comparison showed decrease I₅₀ in iTBS(SC) at FU1 compared (p = 0.002).

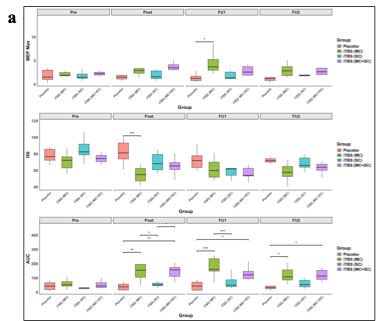

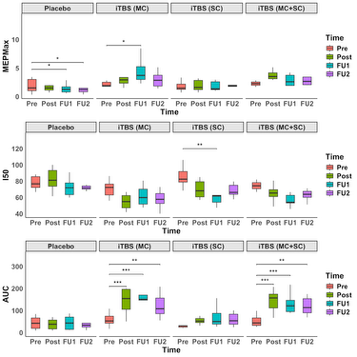

**Supplementary Figure_S10**. **MEP recruitment curve parameters**: MEPmax (mV), I_50_ (%) and AUC (data; median with interquartile range)(a) represent between-group comparisons, and (b) depict within-group comparisons *p < 0.05, **p < 0.01, ***p < 0.001.

**Motor Threshold:** As compared to baseline, a significant reduction was observed in iTBS(MC) (Pre–Post: p = 0.0002; Pre–FU1: p < 0.0001; Pre–FU2: p = 0.013) and iTBS(MC+SC) (Pre–Post: p = 0.0002; Pre–FU1: p < 0.0001; Pre–FU2: p < 0.0001).

**Cortical Silent Period (CSP):** Significant reductions were observed in iTBS(MC) (Pre–Post: p = 0.008; Pre–FU1: p = 0.002) and iTBS(MC+SC) (Pre–Post: p < 0.0001; Pre–FU1: p < 0.0001; Pre–FU2: p = 0.001) as compared to baseline CSP.

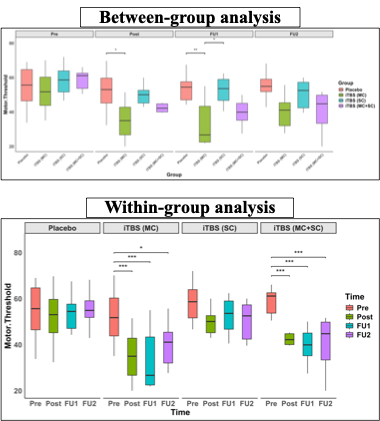

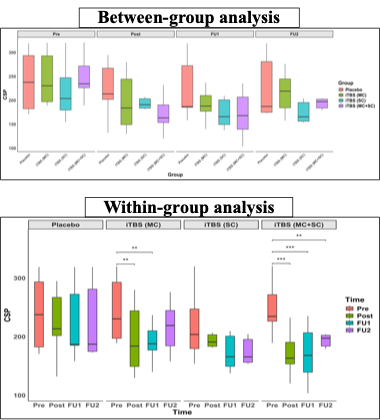

**a**

**b**

### Supplementary Figure_S11. Comparison of (a) Motor Threshold (MT; %) and (b) Cortical Silent Period across stimulation sites (CSP; msec). *p < 0.05, **p < 0.01, ***p < 0.001.

### Short interval intracortical inhibition (SICI)- Intracortical Facilitation (ICF):

### *SICI:* At post intervention, SICI MEP amplitude was significantly higher in the iTBS-SC than in the iTBS-MC group (*p* = 0.0335). By FU2, in all the iTBS-MC, iTBS-SC and iTBS-MC+SC groups significantly higher SICI values were observed than the placebo group (*p* < 0.001, *p* = 0.0015, and *p* < 0.001 respectively). Within-group analysis showed significant increase in the iTBS-MC group (Pre-FU1: *p* = 0.0161 and Pre-FU2: *p* < 0.001) iTBS-MC+SC group (Pre-FU2: *p* < 0.001). In contrast, the iTBS-SC group showed significantly lower SICI values at post-intervention (*p* = 0.0161) and FU1 (*p* = 0.0128) as compared with baseline.

### *ICF*: Between-group comparisons revealed a significantly higher ICF in both the iTBS-MC and iTBS-MC+SC (Post: *p* = 0.0009 and *p* < 0.0001; FU1: *p* = 0.0001 and *p* < 0.0001 respectively) groups as compared to placebo and iTBS-SC (Post: *p* = 0.0034 and *p* = 0.0447; FU1: *p* = 0.0065 and *p* = 0.0023 respectively). Similar findings were observed at FU2, where higher ICF was observed in iTBS-MC and iTBS-MC+SC groups as compared to placebo (*p* < 0.0001) and iTBS-SC (*p* = 0.0004 and *p* = 0.0012, respectively). Within-group analysis showed significant increases from baseline to post-intervention (*p* < 0.0001), FU1 (*p* < 0.0001), and FU2 (*p* = 0.006) in iTBS-MC group. Similarly, the iTBS-MC+SC group exhibited significant increase from baseline to post-intervention, FU1, and FU2 (all *p* < 0.0001).

| **a** | 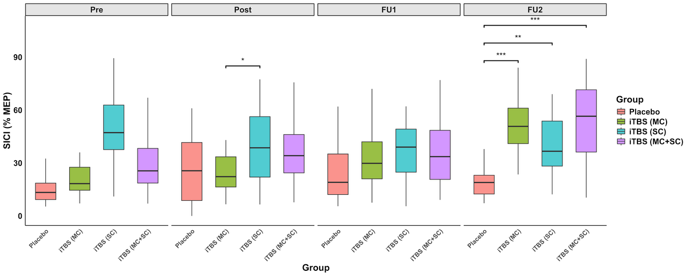 | **b** | 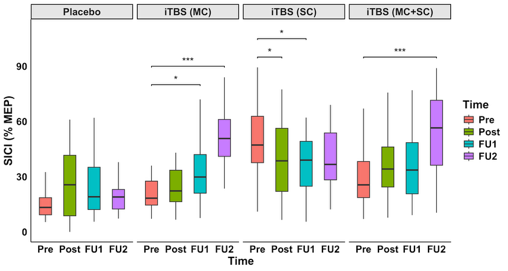 |
| --- | --- | --- | --- |
| **c** | 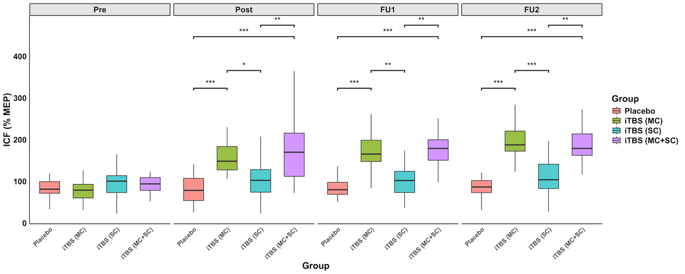 | **d** | 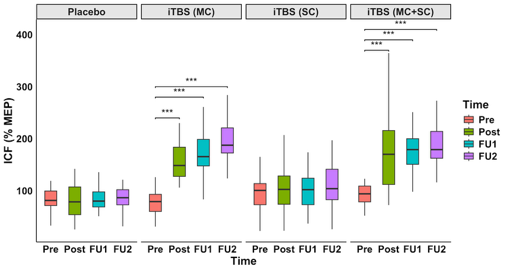 |

### Figure_S12. Short-interval intracortical inhibition (SICI) and Intracortical facilitation (ICF); expressed as %MEP amplitude (Mean ±SEM) across interstimulus intervals at four time points; (a) Between-group comparison of SICI, (b) Within-group changes in SICI, (c) Between-group comparison of ICF and (d) Within-group changes in ICF. Statistical significance is indicated as *p < 0.05, **p < 0.01, ***p < 0.001.

### Long interval intracortical inhibition (LICI): Between-group comparisons showed higher LICI in iTBS-MC as compared to placebo (*p* = 0.0045) and iTBS-SC (*p* = 0.0078) groups at FU2. Even during this period, the iTBS-MC+SC group also showed higher LICI values than the placebo group (*p* = 0.0464). Within-group analysis showed significant increase from baseline to FU1 (*p* = 0.001) and FU2 (*p* < 0.0001) in the iTBS-MC group. A significant increase was also observed between pre-intervention and FU2 (*p* = 0.0034) in the iTBS-MC+SC. In the iTBS-SC group, LICI values were significantly lower at post-intervention (*p* = 0.0066) and FU2 (*p* = 0.017) in comparison to baseline.

| **a** | 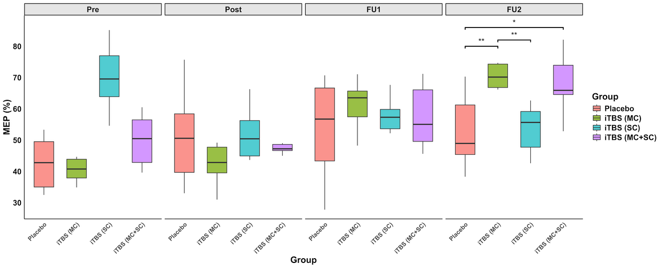 | **b** | 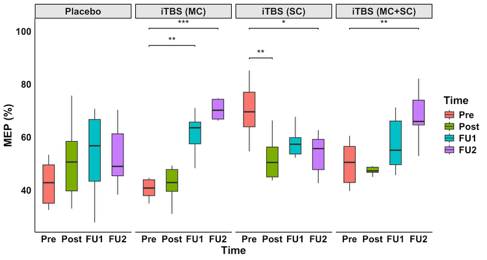 |
| --- | --- | --- | --- |

### Figure_S13. Long-interval intracortical inhibition (LICI); expressed as %MEP amplitude (Mean ±SEM) across interstimulus intervals at four time points; (a) Between-group comparison, (b) Within-group changes. Statistical significance is indicated as *p < 0.05, **p < 0.01, ***p < 0.001.

### Serum biomarkers

**pNF:** At follow-up, in iTBS(MC) the levels were significantly lower than iTBS(SC) (p = 0.032). Within group significant reductions were also observed in iTBS(MC) (at Pre-Post- p = 0.001) and in iTBS (MC+SC) and iTBS (SC) (Pre–FU: p < 0.001 and p< 0.05 respectively).

**MBP:** At Post, MBP was significantly lower in iTBS(MC) relative to placebo (p = 0.044). Significant reductions were also observed in iTBS(MC) at Post (p = 0.011) and follow-up (p = 0.005) as compared to baseline.

**BDNF:** At follow-up, iTBS(MC) and iTBS(MC+SC) was significantly higher than iTBS(SC) (p = 0.003; p = 0.022 respectively). Within-group significant increases were observed in iTBS (MC) only (Pre–FU: p = 0.022).

**NT-3:** Significant increases were observed in iTBS(MC) (Pre–FU: p = 0.022) and iTBS(MC+SC) (Pre–Post: p < 0.0001; Pre–FU: p < 0.0001).

**IL-6:** In comparison to baseline values, a significant reduction was evident at follow up in placebo (p = 0.001), iTBS(MC) (p < 0.0001), and iTBS(MC+SC) (p = 0.0004) groups.

**CD95L:** No significant within-group changes or between-group differences were observed in any group.

**Glutamate:** As compared to baseline, a significant increase were observed in iTBS(MC) (Pre–FU: p = 0.0125) and iTBS(MC+SC) (Pre-Post: p = 0.036) .

**GABA:** Significant reductions were observed in iTBS(MC) (Pre–FU: p < 0.0001) and iTBS(MC+SC) (Pre–FU: p = 0.0003) groups as compared to baseline.

**c**

**b**

**a**

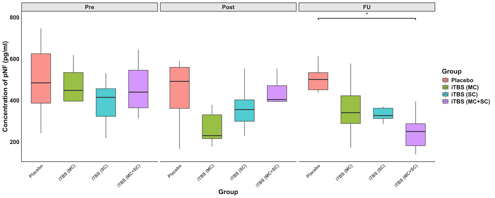

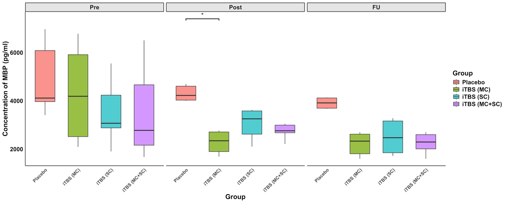

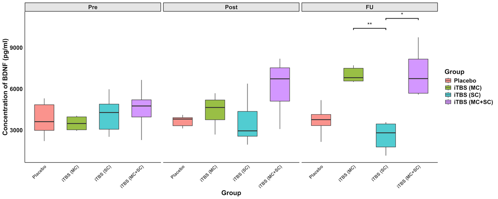

**d**

**f**

**e**

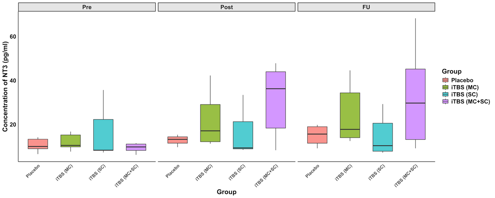

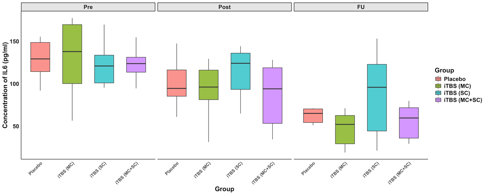

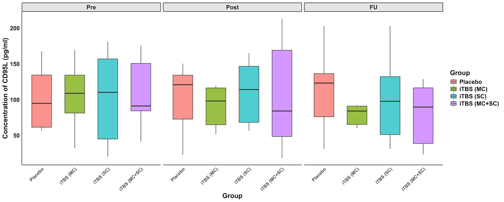

**g**

**h**

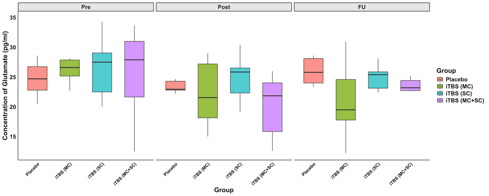

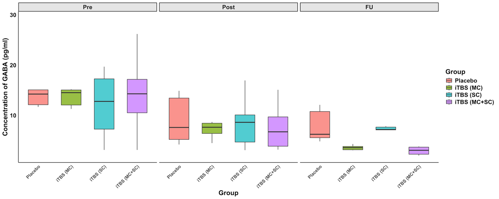

**i**

**k**

**j**

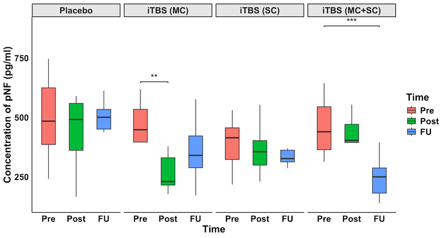

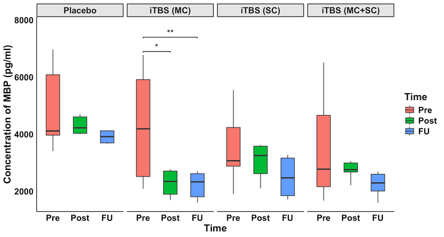

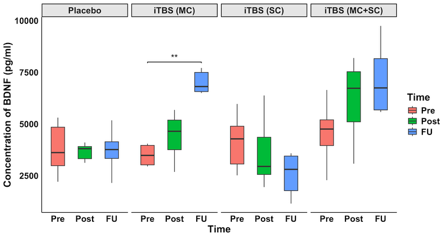

**m**

**l**

**n**

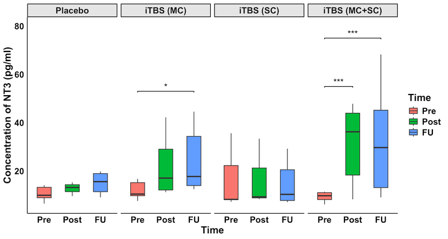

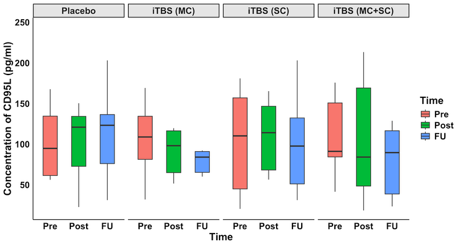

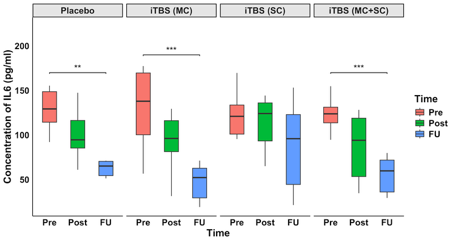

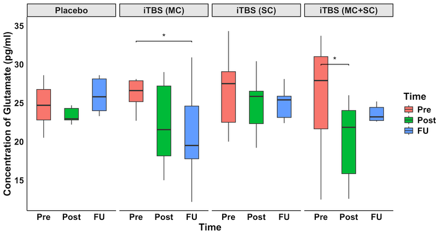

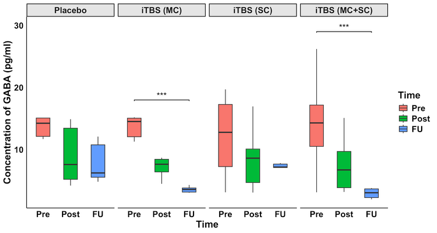

**p**

**o**

**Supplementary Figure_S14. Modulation of serum biomarkers levels across stimulation sites**

**(a–h)** illustrate between-group comparisons (i–p) within-group changes over time for each intervention. Data are presented as boxplots with median and interquartile ranges. *p < 0.05, **p < 0.01, ***p < 0.001. unit: pg/ml for MBP, pNF, BDNF, NT3, IL6 CD95l, GABA and Glutamate).

### Psychosocial outcomes

**WHO-Quality of Life (WHOQOL) domains**

**Physical Domain** score increase significantly in iTBS(MC) (p = 0.030) and iTBS(MC+SC) (p = 0.020) at FU2 as compared to placebo. In comparison to baseline a significant improvement was observed in iTBS(MC+SC) at FU2 (Pre–FU2: p = 0.004). **Psychological Domain** score increase significantly in iTBS(SC) and iTBS(MC+SC) (p = 0.047; p = 0.009) at FU1 and iTBS(MC) and iTBS(MC+SC) (p = 0.014, p = 0.0001) at FU2 as compared to placebo. Within group analysis showed significant improvements in all active groups — iTBS(MC) (Pre–FU1 and Pre–FU2: both p < 0.01), iTBS(SC) (Pre–FU1: p = 0.007; Pre–FU2: p = 0.010), and iTBS(MC+SC) (Pre–FU1: p = 0.0001; Pre–FU2: p < 0.001). In **Social Domain,** scores were found to be higher in iTBS(MC) (Pre–FU2: p = 0.023) and iTBS(MC+SC) (Pre–FU2: p = 0.010) as compared to baseline. **Environmental Domain** showed significant within-group improvement in iTBS(MC+SC) (Pre–FU2: p = 0.0002).

**Beck Depression Inventory (BDI) within group** scores reduced significantly over time in iTBS(MC) (Pre–FU2: p = 0.038), iTBS(SC) (Pre–Post: p = 0.013; Pre–FU1: p = 0.0002; Pre–FU2: p = 0.0002), and iTBS(MC+SC) (Pre–FU2: p = 0.008). **Visual Analogue Scale for Pain (VAS) showed** significant reduction in iTBS(MC) (Pre–FU1: p = 0.030; Pre–FU2: p = 0.004), iTBS(SC) (Pre–Post: p = 0.001; Pre–FU1: p = 0.011; Pre–FU2: p = 0.0004), and iTBS(MC+SC) (Pre–FU1: p = 0.011; Pre–FU2: p = 0.030) groups as compared to baseline. **State and Trait Anxiety (STAI-Y1 and STAI-Y2)** showed no significant within-group or between-group differences.

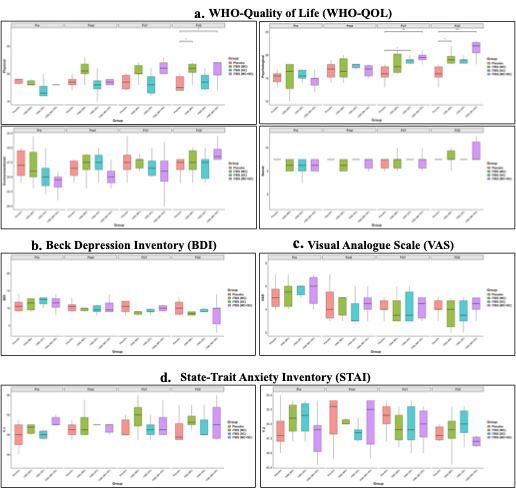

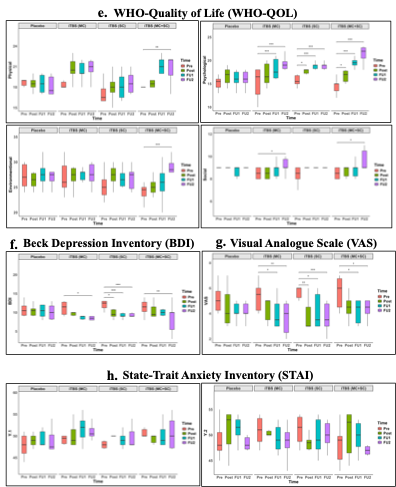

**Supplementary Figure_S15. Psychosocial outcomes in various groups**

(a–d) between-group comparisons, (e–h) within-group changes over time for psychosocial outcomes across four time points: pre-intervention (Pre), post-intervention (Post), first follow-up (FU1), and second follow-up (FU2), across groups (Placebo, iTBS (MC), iTBS (SC), iTBS (MC+SC)). Panel (a, e) represents WHO-Quality of Life (WHO-QOL) domains (Physical, Psychological, Social, Environmental), (b, f) represent Beck Depression Inventory (BDI), (c, g) represent Visual Analogue Scale (VAS), and (d, h) represent State–Trait Anxiety Inventory (STAI-Y1 and STAI-Y2). Data are presented as boxplots showing median and interquartile ranges. Significant differences are indicated (*p < 0.05, **p < 0.01, ***p < 0.001).

**Correlation Analysis**

ΔLEMS was significantly positively correlated with ΔWISCI (R = 0.60, p = 0.002), ΔMEP amplitude (R = 0.49, p = 0.014), ΔAUC (R = 0.43, p = 0.034), and negatively correlated with Δ pNF (R = −0.54, p = 0.006). ΔWISCI were correlated with Δ Mobility (R = 0.79, p < 0.0001), ΔMEP amplitude (R = 0.76, p < 0.0001), ΔAUC (R = .76, p < 0.0001), ΔICF (R = 0.6, p = 0.0021), ΔSICI (R = 0.41, p = 0.044), ΔWISCI showed a significant negative correlation with ΔpNF-H (R = −0.67, *p* = 0.00033), Δglutamate (R = −0.42, *p* = 0.043) and ΔGABA (R = −0.65, *p* = 0.00054).

a.

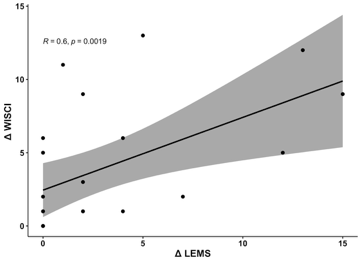

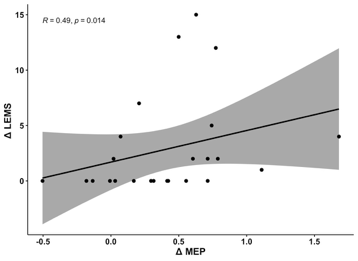

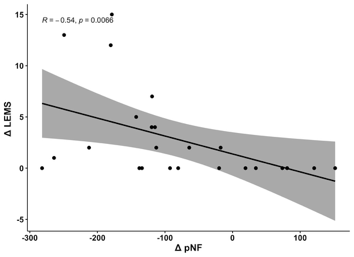

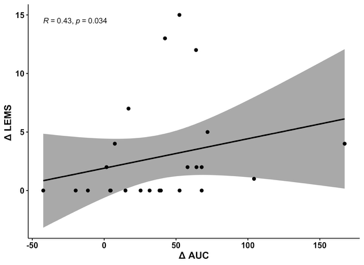

d.

c.

b.

e.

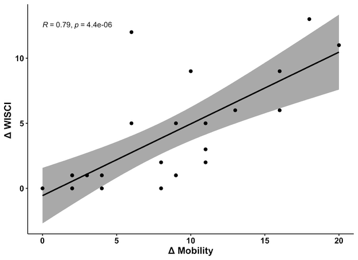

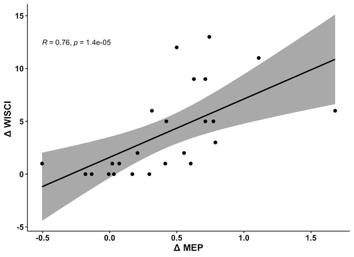

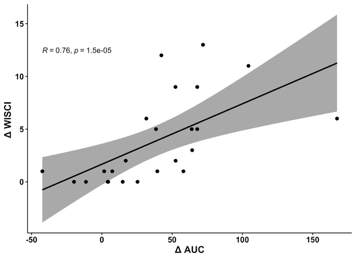

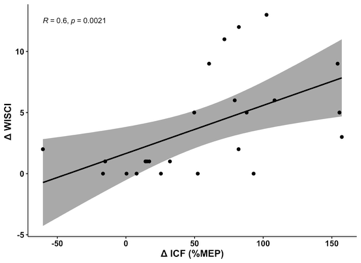

f.

h.

g.

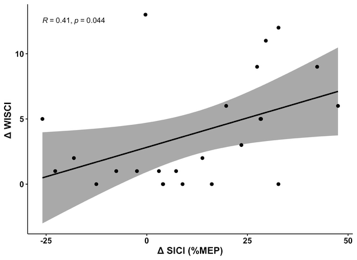

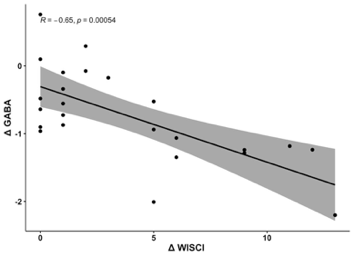

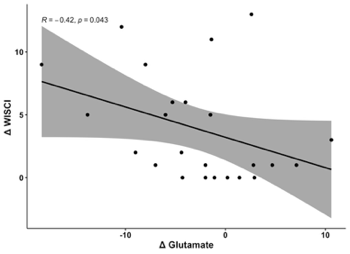

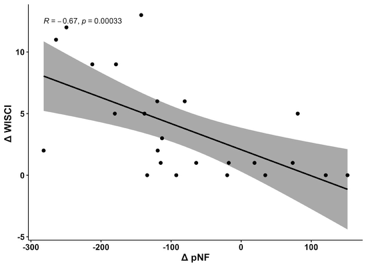

l.

i.

j.

k.

**Supplementary Figure_S16. Spearman correlation analysis between change scores (Δ) in neurological, functional, electrophysiological, and biochemical outcomes following intervention.** **(a–d)** ΔLEMS correlated positively with ΔWISCI, ΔMEP amplitude, and ΔAUC, and negatively with ΔpNF. **(e–l)** ΔWISCI correlated positively with ΔMobility, ΔMEP amplitude, ΔAUC, ΔICF, ΔSICI, and negatively with ΔGABA, Δglutamate and ΔpNF. Shaded bands represent 95% confidence intervals. R = Spearman's rank correlation coefficient, p = p-value, indicating the statistical significance of the correlation (*p < 0.05, **p < 0.01, ***p < 0.001). LEMS = Lower Extremity Motor Score; WISCI = Walking Index for Spinal Cord Injury; MEP = motor evoked potential; AUC = area under the curve; SICI = short-interval intracortical inhibition; ICF = intracortical facilitation; GABA = gamma-aminobutyric acid; pNF = phosphorylated neurofilament.
